# Human SARS-CoV-2 challenge resolves local and systemic response dynamics

**DOI:** 10.1101/2023.04.13.23288227

**Authors:** Rik G.H. Lindeboom, Kaylee B. Worlock, Lisa M. Dratva, Masahiro Yoshida, David Scobie, Helen R. Wagstaffe, Laura Richardson, Anna Wilbrey-Clark, Josephine L. Barnes, Krzysztof Polanski, Jessica Allen-Hyttinen, Puja Mehta, Dinithi Sumanaweera, Jacqueline Boccacino, Waradon Sungnak, Ni Huang, Lira Mamanova, Rakesh Kapuge, Liam Bolt, Elena Prigmore, Ben Killingley, Mariya Kalinova, Maria Mayer, Alison Boyers, Alex Mann, Vitor Teixeira, Sam M. Janes, Rachel C. Chambers, Muzlifah Haniffa, Andrew Catchpole, Robert Heyderman, Mahdad Noursadeghi, Benny Chain, Andreas Mayer, Kerstin B. Meyer, Christopher Chiu, Marko Z. Nikolić, Sarah A. Teichmann

## Abstract

The COVID-19 pandemic is an ongoing global health threat, yet our understanding of the cellular disease dynamics remains limited. In our unique COVID-19 human challenge study we used single cell genomics of nasopharyngeal swabs and blood to temporally resolve abortive, transient and sustained infections in 16 seronegative individuals challenged with preAlpha-SARS-CoV-2. Our analyses revealed rapid changes in cell type proportions and dozens of highly dynamic cellular response states in epithelial and immune cells associated with specific timepoints or infection status. We observed that the interferon response in blood precedes the nasopharynx, and that nasopharyngeal immune infiltration occurred early in transient but later in sustained infection, and thus correlated with preventing sustained infection. Ciliated cells showed an acute response phase, upregulated MHC class II while infected, and were most permissive for viral replication, whilst nasal T cells and macrophages were infected non-productively. We resolve 54 T cell states, including acutely activated T cells that clonally expanded while carrying convergent SARS-CoV-2 motifs. Our novel computational pipeline (Cell2TCR) identifies activated antigen-responding clonotype groups and motifs in any dataset. Together, we show that our detailed time series data (<u>covid19cellatlas.org</u>) can serve as a “Rosetta stone” for the epithelial and immune cell responses, and reveals early dynamic responses associated with protection from infection.

## Main

Coronavirus Disease 2019 (COVID-19) is a potentially fatal disease caused by the severe acute respiratory syndrome coronavirus 2 (SARS-CoV-2), which gave rise to one of the most severe global public health emergencies in recent history. Studies by us and others have uncovered that perturbed antiviral and immune responses to SARS-CoV-2 underlie severe and fatal outcomes, where for example impaired type I interferon responses^1, 2^, decreases of circulating T cell and monocyte subsets^3–5^, and increased clonal expansion of T and B cells^4^ are associated with a more severe outcome. However, accurate detection and interpretation of the immune response during COVD-19 has been hampered by heterogeneous responses caused by numerous non-host factors that affect immune and clinical outcomes that are frequently unmeasurable and uncontrolled. These include infection characteristics such as viral dose, strain and time since exposure, together with clinical features including comorbidities, standard of care and pre-existing immunity. In particular, the observed immune response may represent different phases, from early viral detection to later adaptive responses, depending on the time between infection and sampling.

Since the exact time at which patients were exposed to SARS-CoV-2 is nearly always unknown, it can be challenging to accurately delineate severity-associated and temporal effects such as early interferon signaling and late adaptive immune responses^1–6^. Determining the dynamics of SARS-CoV-2 infection and the body’s response is therefore crucial to understand how the immune response is orchestrated and how risk factors can impact this. In addition, while many studies have investigated responses to COVID-19 during the course of the disease^7, 8^, it has thus far not been possible to study the early phases of exposure and the infection event itself in humans. In particular, studies of natural infection are unable to capture events in those who are exposed to the virus but do not develop sustained viral infection, which might be critical in preventing dissemination and disease. Furthermore, the activation and expansion of antigen-responding T cells (versus bystanders) has been difficult to pinpoint in previous “snapshot” datasets^4, 5^. Here, we trace their development and integrate the paired TCR chains for the first time.

### Human SARS-CoV-2 challenge model

To resolve epithelial and immune cell responses over time from SARS-CoV-2 exposure, we conducted a first of its kind, human COVID-19 challenge study^6^. In this model, young adults seronegative for SARS-CoV-2 spike were inoculated intranasally with a wild-type pre-Alpha SARS-CoV-2 virus strain (SARS-CoV-2/human/GBR/484861/2020) in a controlled environment^6^. Prior to challenge, volunteers underwent extensive screening to exclude risk factors for severe disease and eliminate confounding effects of comorbidities. As risk mitigation and to maximize physiological relevance, participants were inoculated with the lowest culture-quantifiable inoculum dose of 10 Tissue Culture Infectious Dose 50 (TCID_50_). There were no serious adverse events and symptoms resolved spontaneously without treatment.

We studied local and systemic immune responses at single cell resolution in 16 participants. The highly controlled nature of this experimental model allowed baseline measurements on the day before inoculation, followed by detailed time series analyses of cellular responses after inoculation and subsequent infection, both systemically and in the nasopharynx, to decipher antiviral responses against SARS-CoV-2 in a precise time-resolved manner.

Following inoculation, 6 participants from the cohort developed a sustained infection as defined by at least 2 consecutive quantifiable viral load detections by PCR, along with symptoms (**Fig. 1a and Extended Data Fig. 1**). In contrast, three individuals produced multiple sporadic and borderline-positive PCR tests between day 1.5 and 7 post-inoculation. While these participants remained symptom free and did not meet the earlier established criteria to be classified as “sustained infection”, we assigned them to a separate group of “transient infection” to investigate factors associated with this unique phenotype.

**Figure 1:**
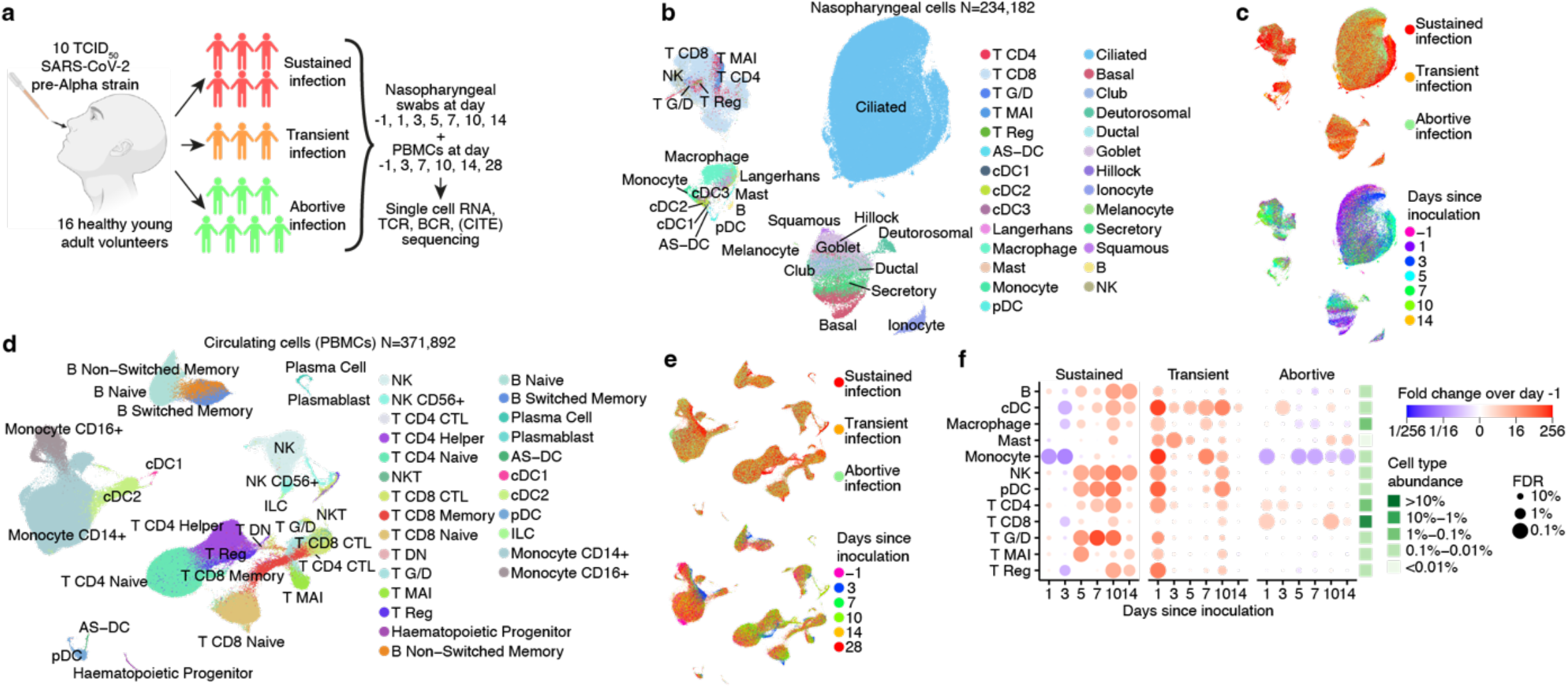
Extensive temporal cell state dynamics after SARS-CoV-2 inoculation. (**a**) Illustration of study design and cohort composition. (**b-c**) UMAPs of all nasopharyngeal cells, color-coded by their broad cell type annotation in (b), by the infection group in the top panel of (c), and by days since inoculation in the bottom panel of (c). Only cells from sustained infection cases are shown in the bottom panel of (c). (**d-e**) UMAPs as in (b-c), but showing all PBMCs. (**f**) Fold changes in abundance of nasopharynx resident broad immune cell type categories. Immune cell abundances were scaled to the total amount of detected epithelial cells in every sample prior to calculating the fold changes over days since inoculation compared to pre-infection (day -1) by fitting a GLMM on scaled abundances. The mean cell type proportions over all cells and samples is shown in the green heatmap right of the dotplot to aid the interpretation of changes in cell type abundances.

Seven participants remained PCR negative throughout the quarantine period, indicating that these individuals successfully prevented the onset of a sustained or transient infection. Due to the fact that these participants all remained seronegative, but were observed to display early innate immune responses (see below), we termed these abortive infections (as opposed to uninfected due to for example antibody-mediated sterilizing immunity).

### Broad cellular transitions observed over time and infection groups

To comprehensively identify and time responses to SARS-CoV-2 exposure in these phenotypically-divergent groups, we performed single cell RNA sequencing (scRNAseq) and single cell TCR- and BCR-seq at up to seven time points (**Fig. 1a**). In addition, we complemented the RNA measurements in PBMCs with CITE-seq measurements to quantify 123 surface proteins to aid cell type annotation. At each time point, we collected PBMCs and nasopharyngeal swabs to study both the systemic immune response and the epithelial and local immune response at the site of inoculation, respectively. Of note, while most PBMC and nasopharyngeal time points were matched, we included more early nasopharyngeal and later PBMC time points as we anticipated more immediate local responses. In total, we generated over 600K single cell transcriptomes across 181 samples, which include 371,892 PBMCs and 234,182 nasopharyngeal cells. We used predictive models and marker gene expression to annotate 202 cell states in total (see *Methods*; **Extended Data Fig. 2-5**), including multiple newly identified cell states that will be discussed throughout this manuscript. Importantly, both datasets contained all expected cell types (**Fig. 1b,d; Extended Data Fig. 2a-b**), including clearly resolved epithelial and immune compartments in the nasopharyngeal samples, which enabled us to study both the local and systemic immune response. Strikingly, even when visualizing all cells at once in **Fig. 1b,d**, the “infection group” and “days since inoculation” mark specific groups of cells (**Fig. 1c,e**), indicating that there are large changes in cell fate over time and infection groups across the different cell type compartments.

### Innate and adaptive immune infiltration to site of inoculation

We first investigated how the immune landscape is affected by viral inoculation and subsequent infection. We used generalized linear mixed models (GLMM) to quantify the changes in cell type abundances over time since inoculation compared to the day prior to inoculation (-1). This allowed us to perform paired longitudinal modeling of donor-specific effects while accounting for technical and biological variation using random effect terms. Analysis of the nasopharyngeal resident immune compartment revealed that all immune cell types significantly infiltrate the site of inoculation after exposure to SARS-CoV-2 (**Fig. 1f**). Strikingly, the timing of infiltration strongly differed between participants with a sustained, transient or abortive infection. During sustained infections, immune infiltration started at day 5 after inoculation and continued to increase until day 10. In stark contrast, transient infections led to immediate and robust immune infiltration, followed by a decrease and smaller secondary infiltration event at day 10. Last, abortive infections do not lead to any obvious patterns of immune infiltration.

Interestingly, both sustained and transient infections lead to infiltration of innate and adaptive immune cells. However, the increase of innate immune cells such as plasmacytoid dendritic (pDC), natural killer (NK), gamma/delta T (g/d T), and mucosal-associated invariant T (MAIT) cells was quicker and of greater magnitude than infiltration by adaptive immune cells in sustained infections. In line with this, in transient infections, the increase of immune cells at day 1 was also greatest in the innate immune compartment. The observed difference in timing of immune infiltration between transient and sustained infections suggests that immediate immune recruitment and responses are associated with containing SARS-CoV-2 infection and preventing the onset of a sustained infection and COVID-19.

### Widespread systemic interferon response precedes response at site of inoculation

We next attempted to detect antiviral gene expression programs in any of the tissue resident and circulating cells during infection. Gene expression analysis revealed that the interferon response genes made up the dominant infection-induced gene expression module in participants with sustained infection (**Fig. 2a**). Strikingly, interferon signaling was strongly activated in every cell type of both the blood and in the nasopharynx, where up to 100% of some cell types at a given time took on a distinct interferon stimulated cell state (**Extended Data Fig. 6a**, annotated in **Extended Data Fig. 2-4** as IFN stim), underscoring its widespread and dominant effect. Activation of interferon signaling was absent in abortive infection and only short-lived in transient infections (**Extended Data Fig. 6a**). Interestingly, at the site of inoculation, we only detected widespread interferon activation from five days post-inoculation, whereas the interferon response in the blood peaks at day 3 post-inoculation and appears to be stronger. This is unexpected, as we assumed that the cells that reside in the inoculated tissue should be the first to respond through direct exposure to the virus and infected cells. Instead, it appears that the immediate response to SARS-CoV-2 infection includes informing circulating immune cells before tissue-resident cells through interferon signaling.

**Figure 2:**
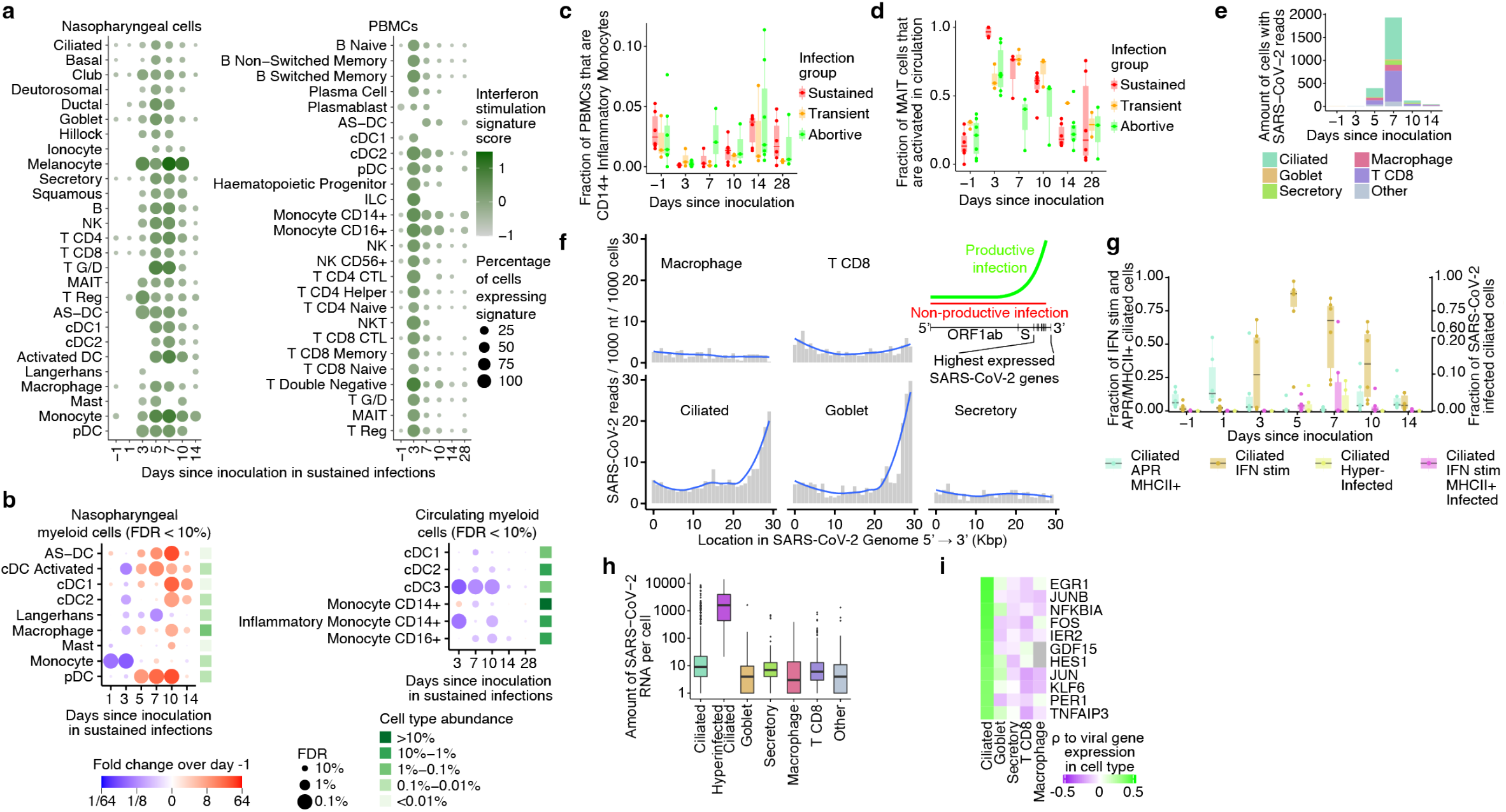
Cell-state-specific antiviral responses and infection. (**a**) Dotplot visualizing the mean expression of interferon stimulated genes across cell types and time since inoculation in participants with sustained infections, for nasopharyngeal cells (left plot) and PBMCs (right plot). (**b**) Dotplot as in (Fig 1f), showing myeloid cell types in sustained infection cases that significantly change at least one time point compared to pre-infection. Nasopharyngeal cells and PBMCs are shown in the left and right plot, respectively. (**c**) Boxplot showing the relative amounts of circulating inflammatory monocytes over time since inoculation in each infection group. (**d**) Boxplot showing the fraction of circulating MAIT cells that are activated over time since inoculation in each infection group. (**e**) Stacked barplot showing the amount of nasopharyngeal cells with at least one SARS-CoV-2 RNA read detected (after background subtraction), split by days since inoculation and color-coded by cell type. (**f**) Barplots showing the distribution of detected viral reads over the SARS-CoV-2 genome in the five most highly infected cell types. The blue line represents a loess fit over the data. The top-right inset illustration is shown to aid the interpretation of a uniform read distribution versus a 3’ biased read distribution. (**g**) Boxplot showing the fraction of ciliated cells that are annotated into detailed response or infection cell states. Only cells from sustained infection cases are shown and split by days since inoculation. The Y axis for interferon (IFN) and acute-phase response (APR) positive ciliated cells is shown on the left, while the Y axis for infected ciliated cells is shown on the right. (**h**) Boxplot of the amount of viral sequencing reads per cell type. (**i**) Heatmap of spearman correlations between host gene expression and the amount of viral reads found in each cell, split by cell type. Shown genes correlate the highest with gene expression in ciliated cells. In all box plots, the central line and the notch are the median and its approximate 95% confidence interval, the box shows the interquartile range and the whiskers are extreme values upon removing outliers.

### Temporal reduction of myeloid subsets immediately after viral exposure

To investigate the potential role of professional antigen presenting cells in the early immune response to SARS-CoV-2, we next focused on changes in the nasopharyngeal resident and circulating myeloid compartments during sustained infection. In contrast to most tissue-resident immune cells, myeloid subsets largely decreased in frequency at the site of inoculation by day 3 after inoculation (**Fig. 2b**). In particular, monocytes and activated DCs (also often referred to as migratory DCs, mature DCs or DC-LAMPs) were significantly reduced at the site of inoculation at day 3, consistent with the known role of activated DCs in trafficking viral antigens to lymph nodes for presentation. After day 3, the number of DCs at the site of inoculation increased above baseline, along with the global immune infiltration associated with peak viral load in sustained infections. We also observed a decrease of circulating myeloid cells during infection across all subsets at day 7, consistent with continued migration of these cells into inflamed or lymphoid tissues at that time point. However, already at day 3 post-inoculation there are strong decreases in some circulating myeloid subsets, where inflammatory monocytes (*IL1B, IL6* & *CXCL3* high) and cDC3s (monocyte-like dendritic cells) appear to migrate out of the circulation.

Strikingly, while all of the above mentioned responses were only observed in sustained infections, the significant decrease in inflammatory monocytes was also observed in transient and abortive infections (**Fig. 2c** and **Extended Data Fig. 7a**). This suggests that circulating inflammatory monocytes are able to immediately respond to SARS-CoV-2 exposure, even if the viral infection is rapidly terminated, implying that exposure alone in the absence of virologically-confirmed infection with SARS-CoV-2 can result in a detectable (but restricted) immune response.

### Novel MAIT subset is activated in both sustained and abortive infections

We next asked if such a detectable immune response across all infection groups could also be observed in other cell types. When annotating unconventional T cells, we noted that MAIT cells could be further divided into two subgroups, i.e. classical MAIT cells and activated MAIT cells with higher expression of cytotoxicity and activation markers such as *PRF1* and *CD27* (**Extended Data Fig. 6b**). These markers have previously been shown to be indicative of TCR-independent activation^9^. At day 3 post-inoculation, we observed near complete activation of the entire MAIT cell population in the blood in sustained infections (**Fig. 2d**). Strikingly, the activation of MAIT cells was also present in abortive and transient infections, which suggests that MAIT cells may rapidly sense exposure to a virus. Thus both MAIT cells and inflammatory monocytes might play a key role in the immediate response to SARS-CoV-2. This further supports the notion that viral exposure, that does not lead to a sustained infection and subsequent COVID-19, can still induce a detectable, yet restricted immune response.

### Infection in epithelial and immune cells peaks a week after exposure and is most active in ciliated cells

To study how the observed immune responses relate to viral infection dynamics, we included the SARS-CoV-2 ssRNA genome and its transcripts in our analyses. This allowed us to quantify virions and viral gene expression alongside transcriptome dynamics of infected host cells. As expected, infected cells were almost exclusively found in the nasopharynx of participants with sustained infections (2505 out of 2512 cells with viral RNA). We detected infection of multiple cell types at day 5 post-inoculation, which peaked at day 7 (**Fig. 2e**), followed by a rapid decrease at day 10 - 14 post-inoculation, showing the narrow time window over which SARS-CoV-2 virion production occurred. These changes over time were in line with qPCR results (**Extended Data Fig 1b,c and Extended Data Table 1a,b**), albeit with the latter being more sensitive. Interestingly, we observed viral reads in both immune and epithelial cells in the nasopharynx (**Fig. 2e**). In contrast to previous studies, we detected large numbers of SARS-CoV-2-containing CD8+ T cells, possibly due to the model being able to capture the narrow time window in which these infected cells are highly abundant. Our results therefore show that, in addition to epithelial cells, viral transcripts are also detectable at high levels in tissue-resident CD8+ T cells.

### Non-productive SARS-CoV-2 infection of immune cells

Having identified infected cells, we next asked if these represented productive infections. Because SARS-CoV-2 has a polyadenylated ssRNA genome, we were able to detect both viral transcripts and genomes, which allowed us to separate non-productive and productive infections. In non-productive infections only viral genomes would be present, leading to a fairly uniform distribution of detected viral RNAs over the length of the viral genome (slightly 5’ biased due to 5’ tag sequencing). In contrast, a productive infection requires viral transcription which is known to be highly biased towards the 3’ end of the viral genome^10^. We observed that the viral RNA found in infected ciliated and goblet cells mainly originated from the 3’ end where most genes are encoded, while viral RNA was uniformly distributed across the genome in infected CD8+ T cells and macrophages (**Fig. 2f**). This suggests that immune cells are not permissive for or are capable of preventing viral transcription and subsequent replication after entry into the immune cell, while goblet and ciliated cells are susceptible to proliferative viral infection. While the detection of inactive SARS-CoV-2 in macrophages could be a consequence of the engulfment of virions and infected cells, it is unclear how infection of tissue-resident CD8+ T cells is achieved and how this affects their function.

### Hyper-infected ciliated cells are the main source of SARS-CoV-2 and produce anti-inflammatory molecules

Based on the detection of productive viral infections in ciliated and goblet cells, we sought to identify the cells that contributed the most to viral spread. We noticed a small but distinct cluster of ciliated cells with an extremely high viral load (**Fig. 2h**, **Extended Data Fig. 2b**), in which we detected >1000 viral RNAs per cell on average. Other infected cells typically contained <10 detectable viral RNAs. Strikingly, while this hyper-infected subcluster of ciliated cells represents only 4% of all infected cells, they contained 67% of all detectable viral RNA, uncovering an important role for this subset of ciliated cells in fueling the viral spread.

To investigate how the varying amounts of virus per cell affects host cell gene expression, and *vice versa*, we correlated the amount of viral RNA with the expression of host genes. This revealed that ciliated cells exhibited a unique response to high viral amounts, upregulating AP1 and NFKB signaling, and multiple genes with known anti-inflammatory functions such as ERG1^11^, NFKBIA^12^, GDF15^13^, HES1^14^, PER1^15^, TNFAIP3^16^, and NR4A1^17^ (**Fig. 2i**). This suggests that SARS-CoV-2 is capable of inducing an unique response state in hyper-infected ciliated cells that is in part anti-inflammatory, possibly to enhance viral spread and survival. This is further supported by the attenuation of the interferon response in hyper-infected ciliated cells (**Extended Data Fig. 6c**).

### Ciliated cells exhibit multiple temporally restricted response states resulting in MHC class II presentation on infected cells

To further investigate the role of ciliated cells in the local response to SARS-CoV-2 infection, we delineated the ciliated cell compartment into five distinct cell states. In addition to the above-mentioned interferon-stimulated, infected, and hyper-infected clusters, we detect a relatively abundant subset of ciliated cells with high expression of acute-phase response (APR) genes such as SAA1. Interestingly, these APR+ ciliated cells are present before inoculation, and increase the day after inoculation to up to 50% of all ciliated cells in participants with sustained infections (**Fig. 2g**). At day 3 post-inoculation, interferon-stimulated ciliated cells emerge and peak at day 5, at which time point APR+ ciliated cells have disappeared completely. At day 5, infected and hyper-infected ciliated cells start appearing, which peak at day 7 post-inoculation. At day 10-14, interferon-stimulated cells decrease but remain higher than baseline, while APR+ ciliated cells reemerge. Of note, APR+ ciliated cells are also immediately upregulated in abortive but not transient infections, while all other ciliated cell states are uniquely present in sustained infections only (**Extended Data Fig. 7c**).

Together, this underscores the highly dynamic nature of the ciliated cell compartment, and uncovers a potential early response role for APR+ ciliated cells. Interestingly, infected but not hyper-infected, ciliated cells also activate APR genes, and both APR+ and APR+/SARS-CoV-2 infected ciliated cells express MHC class II (**Extended Data Fig. 6c**). While epithelial cells normally only express MHC class I to present antigens to CD8+ T cells, there is evidence that viral infection can also induce MHC class II expression in epithelial cells^18^, despite classically being thought to be an exclusive feature of professional antigen presenting cells. The colocalization of MHC class II+ ciliated cells with CD4+ T helper cells has also previously been reported^19^. This therefore raises the possibility that MHC class II expression in infected ciliated cells could allow these cells to present SARS-CoV-2 antigens to antigen-specific CD4+ T cells.

### Identification of activated T cells

To investigate the anatomic and temporal distribution of CD4+ and CD8+ T cells following infection, we annotated the T cell compartment in the blood and nasopharynx at high resolution into 54 distinct T cell states (**Fig. 3a,e**). Strikingly, these included subtypes of CD4+, CD8+ and regulatory T cell states that highly expressed T cell activation markers such as *CD38*, *CD28*, *CD27* and *ICOS* (**Fig. 3b**). While we and others have previously been unable to separate T cells that become activated during SARS-CoV-2 infection without enrichment experiments, we detected these activated T cells as distinct clusters in both the circulating and nasopharyngeal T cell compartments. Reassuringly, many nasopharyngeal and circulating activated T cells expressed the same TCR sequences (**Extended Data Fig. 6d**), showing that they originated from the same clones found both in circulation and nasopharynx as a response to infection. In addition, the immune repertoires of activated T cells were significantly more restricted and clonal than other mature T cell types (**Extended Data Fig. 6k**), suggesting that they were activated and expanded in a TCR- and antigen-specific manner. As expected from activation through TCR signaling, we also detected high frequencies of cycling T cells within the activated T cell compartment. Of note, we also detected many activated T cells that were not cycling, as well as cycling T cells that did not appear to be activated, implying that our activation signature was at least partially independent of the cell cycle gene signature. To test if these newly identified activated T cells are antigen-specific and can recognize SARS-CoV-2 peptides, we performed peptide-MHC-I stainings on PBMCs using DNA-barcoded Dextramers loaded with SARS-CoV-2 antigens to detect peptide-MHC-I binding in parallel with scRNAseq and scTCRseq. These experiments reveal that activated T cells are significantly enriched and indeed specifically bind SARS-CoV-2 peptides compared to unmatched peptide-MHC-I molecules (**Extended Data Fig. 7d-f, 8b**). Together, the identification of activated T cells and their transcriptome signature in unsorted PBMC and tissue samples presents a unique opportunity to study the T cell response to SARS-CoV-2 in unprecedented detail.

**Figure 3:**
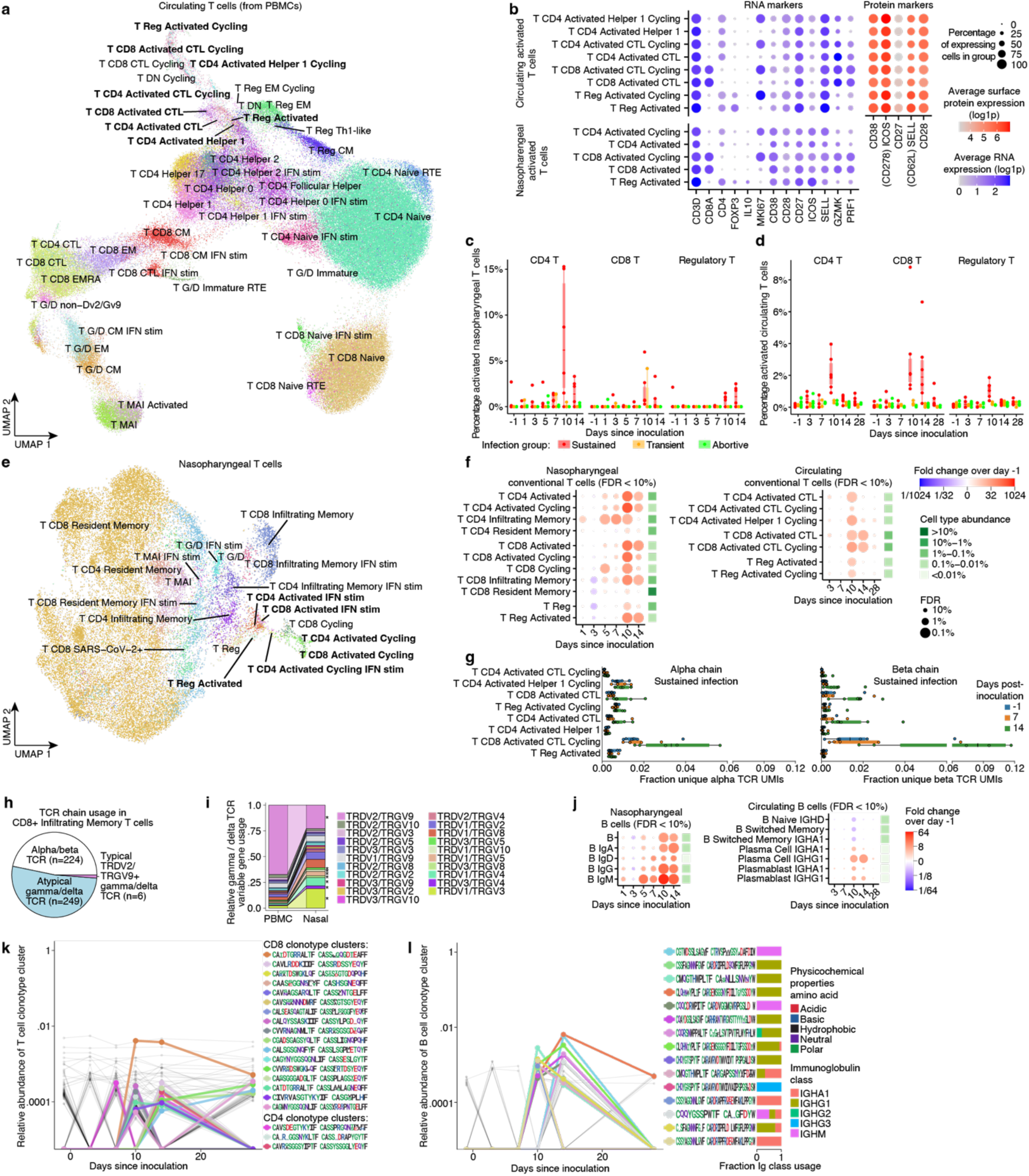
Adaptive immune responses emerge at day 10 post-inoculation. (**a**) UMAP of all circulating T cells, highlighting the distinct cluster of activated T cells. Cells are color coded and labeled by their detailed cell state annotation. (**b**) Marker gene and protein expression of activated T cell subsets are shown in blue and red, respectively. (**c**) Percentages of nasopharyngeal T cells that were annotated as activated T cells, split over days since inoculation and color coded by infection group. (**d**) Boxplot as in (c), but showing circulating activated T cells. (**e**) UMAP as in (a), but showing nasopharyngeal T cells. (**f**) Fold changes in cell state abundance compared to pre-inoculation of nasopharyngeal and circulating conventional T cells are shown in the left and right plots, respectively. Only cell states that significantly change at a FDR < 10% at least one time point are shown. Nasopharyngeal T cell abundances were scaled to the total amount of detected epithelial cells. Fold changes and significance were calculated by fitting a GLMM as shown in Figure 1. The mean cell type proportions over all cells and samples is shown in the green heatmap right of the dotplot to aid the interpretation of changes in cell type abundances. (**g**) TCR clonality and expansion at day 14 of activated TCRs was validated using bulk TCR sequencing. For TCRs that matched the single cell gene expression, normalized clonality TCR alpha (left) and beta (right) data is separated by type and expressed as the average fraction of total clones in sample contributed by a cell of that type, with changes over time implying clonal expansion or contraction. For activated T cell types of interest, scatterplots for each sustained infection and at each time point sampled (days -1, 7, 14) are drawn. (**h**) Proportion of CD8+ infiltrating T cells that use alpha/beta TCRs, typical Dv2/Gv9 g/d TCRs, or atypical g/d TCRs is shown. (**i**) The relative immune repertoire composition of g/d T cells in circulation and nasopharynx after challenge are shown in the left and right bars, respectively. G/d chain pairs that are significantly more or less abundant between circulation and nasopharynx (p<0.05) are highlighted with an asterisks. (**j**) Dotplot as in (f), showing the fold changes in B cells. Legend for significance and mean cell type proportions as in (f). (**k**) Abundance of TCR clusters relative to all TCRs are shown over time since inoculation. Activated TCR clusters are color coded and their TCR motifs are shown. Legend for the physicochemical properties of amino acids in shown TCR motifs is shown in panel (l). (**l**) Plot as in (k), but showing BCR clusters. Immunoglobulin class usage within each activated BCR cluster is shown in the rightmost bars.

### Activated T cells expand and peak ten days after inoculation

To better understand the characteristics of the activated T cells described above we quantified their abundance over time and across infection groups (**Fig. 3c-d,f**). This revealed highly significant expansions of activated CD4+ and CD8+ T cells peaking in both blood and nasopharynx at day 10 after inoculation. This expansion was highly time-restricted, only appearing in the circulation after day 7 and contracting rapidly thereafter. While this decrease meant that activated T cells were barely detectable at day 28 post-inoculation, the associated TCR clonotypes in circulation could still be identified, having transitioned into memory and effector T cells (**Extended Data Fig. 6e**). We integrated our single cell resolved T cell data with highly sensitive bulk TCR sequencing from the blood to validate that activated T cell-associated TCR sequences indeed clonally expand after day 7 post-inoculation in sustained (**Fig. 3g**) but not in abortive infections (**Extended Data Fig. 6f**). The emergence of these cells at day 10 after inoculation closely resemble the temporal dynamics of a typical antigen-specific adaptive immune response to vaccination and infection. At this time point we also observed clearance of detectable virus and a reduction of IFN stimulation in the nasopharynx, suggesting that the onset of an adaptive T cell response is associated with clearance of the infection. Importantly, activated T cells emerged in all sustained infected participants, but in none of the abortive infections, underscoring their specificity to infection. We did however detect a small increase of activated T cells in the nasopharynx of two of the three transiently infected individuals (**Fig. 3c**), which might suggest that a smaller T cell response can be established without going through a sustained infection.

In contrast to activated CD4+ and CD8+ T cells whose infiltration peaked at day 10, the amount of activated regulatory T cells was highest at day 14 at the site of infection (**Fig. 3c**), where they strongly upregulated expression of the anti-inflammatory cytokine IL-10 (**Fig. 3b**). This peak of activated regulatory T cells coincided with resolution of the observed global immune infiltrate (**Fig. 1f**) and downregulation of the IFN-stimulated response (**Fig. 2a**), suggesting a role for these regulatory T cells in suppressing further local inflammation after the infection has been cleared.

Interestingly, the time window during which activated CD8+ T cells were increased was broader in blood (**Fig. 3d**), while activated CD4+ T cells were detected for longer in the nasopharynx (**Fig. 3c**). In addition, activated CD4+ T cells were also significantly more abundant at the site of infection where they represent up to 15% of all nasopharyngeal-resident T cells at day 10 after inoculation. The predominance of activated CD4+ T cells in the respiratory mucosa was surprising, as CD8+ T cells are classically understood to be the major effectors in the local cytotoxic response. These results suggest that CD4+ T cells may play an unexpected and important role as local effectors.

### Activated CD4+ T cells express cytolytic proteins

Activated CD4+ T cells express high amounts of cytotoxicity genes (e.g. *PRF1*, see **Fig. 3b** and **Extended Data Fig. 3a & 4a)** that are normally expressed in NK and CD8+ T cells. As CD4+ T cells can only recognise antigens in MHC class II context that is normally exclusive to professional antigen-presenting immune cells, the function and relevance of cytotoxic CD4+ T cells remains poorly understood. However, several studies have reported their emergence during the adaptive immune response against SARS-CoV-2^20, 21^, and they have been reported to have a specific and antiviral effector function in influenza challenge models^22^. It is therefore conceivable that the activation of cytotoxic CD4+ T cells upon SARS-CoV-2 infection could potentially be a response to MHC class II presentation by infected ciliated cells (**Extended Data Fig. 6c**). This would potentially enable antigen-specific destruction of infected ciliated cells by CD4+ T cells (**Fig. 2g**).

### Atypical g/d T cells infiltrate site of infection and dominate the g/d T cell response

In the nasopharynx, we also detected a subset of CD4+ and CD8+ T cells lacking both activation and tissue-residency markers (such as *ITGAE* or CD103, **Extended Data Fig. 3a**) which appeared in the nasopharynx during sustained infections and which were annotated as “infiltrating memory T cells” (**Fig. 3e**). We noticed that infiltrating CD8+ T cells expressed relatively few detectable alpha/beta TCRs and had heterogenous CD8 expression, similarly to the gamma/delta (g/d) T cells that we had already detected (**Extended Data Fig. 2a**). To investigate if this infiltrating subset harbors a distinct g/d T cell population, we performed targeted single cell sequencing of the g/d TCR genes in the nasopharynx and blood. This revealed that infiltrating CD8+ T cells indeed predominantly express g/d TCRs (**Fig. 3h**). As expected, the g/d TCR repertoire found in circulating blood cells consists mostly of TCR chains containing variable segments TRDV2 and TRGV9 (**Fig. 3i**). Strikingly, the nasopharynx is significantly depleted for TRDV2/TRGV9+ T cells, with other variable segments dominating the g/d TCR repertoire. More than 97% of the g/dTCR expressing infiltrating CD8+ T cells express these rare non-TRDV2/TRGV9 TCRs (which we termed atypical g/d T cells; **Fig. 3h**), which means that the atypical g/d T cell response is four times more abundant than the typical g/d T cell response. While the exact function of atypical g/d T cells is still poorly understood, their timing alongside other adaptive immune responses and its restriction to sustained infections, suggests that they might play an underappreciated role in the immune response against SARS-CoV-2 infection.

### Antibody secreting B cells clonally expand ten days after exposure

Given that the strong T cell response that appears highly time restricted to day 10 post-inoculation, we hypothesized that there should be a B cell response at a similar time point. To test this, we investigated the temporal and cell state dynamics of the B cell response to SARS-CoV-2 inoculation. We detected distinct subtypes of naive, memory and antibody-secreting B cells (plasmablasts and plasma cells), and used the BCR data to distinguish immunoglobulin class and isotype switching (**Extended Data Fig. 4b**). In line with the observed T cell response, we observe a strong and highly time restricted B cell response from day 10-14 after SARS-CoV-2 exposure (**Fig. 3j**). In blood, this response includes a clear switch from naive and IgG/IgA memory B cells to mostly IgG1 and some IgA1 secreting plasmablasts and plasma cells. IgA1 and IgG1 are expected to be the dominant antibody immunoglobulin classes in blood^23^, and the timing of production of antibodies is in line with B cell responses observed in vaccination studies^24^, suggesting that these antibody secreting B cells at day 10 after inoculation are SARS-CoV-2 specific. While numbers of detected B cells in the nasopharynx are limited, we also observe significant infiltration of both IgA+ and IgG+ B cells into the nasopharynx from day 10 post-inoculation (**Fig. 3j**), indicating that the B cell response leads to antibody production at the site of infection. Together, these findings suggest that it takes ten days from SARS-CoV-2 exposure for the adaptive immune response to mature and expand to detectable abundances. Importantly, we show a concerted adaptive immune response of B and T cells at both local and systemic level, which is facilitated by antibody secreting and activated lymphocytes.

### Cell2TCR: Identification of clonotype groups that are likely SARS-CoV-2 specific

We next set out to leverage the transcriptomically distinct B and T cell states that are associated with the adaptive immune response, to identify BCR and TCR clonotypes that specifically recognise SARS-CoV-2 (see *Methods* for details). We designed a cell state-driven approach that enabled us to detect the permitted divergence between TCR or BCR sequences in an antigen-specific response. To this end, we quantified the inclusion of naive B and T cells, and the mixing of CD8+ and CD4+ T cells, to quantify the impurity of TCR and BCR clonotype groups (groups of B and T cells that express highly similar BCR or TCR sequences). We next selected activated clonotype groups that appear to expand in an antigen-specific manner (i.e. express multiple independent but highly similar TCR/BCR sequences in activated T cells or antibody-secreting B cells). Reassuringly, this clonotype selection method exclusively yields activated clonotypes in participants with sustained infections (**Extended Data Fig. 6g-h**). In total, we detect 20 activated TCR clonotype groups and 15 activated BCR clonotype groups in the six participants with sustained infections (**Fig. 3l**). These clonotype groups first emerge after 1 week, most appear at day 10, and some remain detectable at day 28 post-inoculation. When we applied Cell2TCR on all activated CD8+ T cells as well as all HLA-matched CD8+ T cells from the Dextramer assay, we found 14 clonotype groups that contained cells from both datasets, validating the specificity and annotating the recognized antigen of these clonotype groups (**Extended Data Table 1c**).

Interestingly, even at the peak of expansion at day 10 post-inoculation, all but one of the activated clonotype groups have only very low abundance (<0.001% of all T cells), at the detection limit of single cell genomics approaches. Such low prevalence makes activated clonotypes difficult to detect and distinguish from bystander cells when simply performing enrichment analysis of the entire immune repertoire between healthy and infection samples. This highlights the importance of considering single cell phenotypes in VDJ analyses and the usefulness of our newly identified activated T cell state.

Importantly, in contrast to activation or enrichment assays that require *in vitro* incubation with antigens^25, 26^, our Cell2TCR approach for detecting clonotypes that are activated in a disease of interest is not restricted and biased towards known antigens. Hence, it can be applied to any infection, inflammatory disease or cancer single cell RNA- and VDJ-seq dataset to extract paired chains recognising antigens.

### Integrating COVID-19 patient data uncovers public SARS-CoV-2 TCR motifs

We hypothesized that our deep characterization of the adaptive immune response in PBMCs could be leveraged when analyzing patient COVID-19 data, in particular to study activated T cell states and associated SARS-CoV-2 specific TCR repertoires. To this end, we integrated our data together with single cell RNA-seq data from five large-scale studies that profiled PBMC samples using a deep generative model (scVI variational autoencoder, see *Methods*), and obtained just short of one million T cells from several hundred individuals, including over 240 acute COVID-19 patients (**Extended Data Table 1d,e**). We next projected our highly detailed cell type annotation, including the activated T cell states, onto the patient data (**Fig. 4a and 4c**). This revealed that activated T cells are also present in COVID-19 patients that were sampled outside a viral challenge setting, and that these activated subsets also form distinct clusters of cells within the T cell compartment. Importantly, the fraction of activated T cells was significantly higher in COVID-19 patients and convalescence samples compared to healthy controls, underscoring their involvement in the immune response to COVID-19 (**Fig. 4b**).

**Figure 4:**
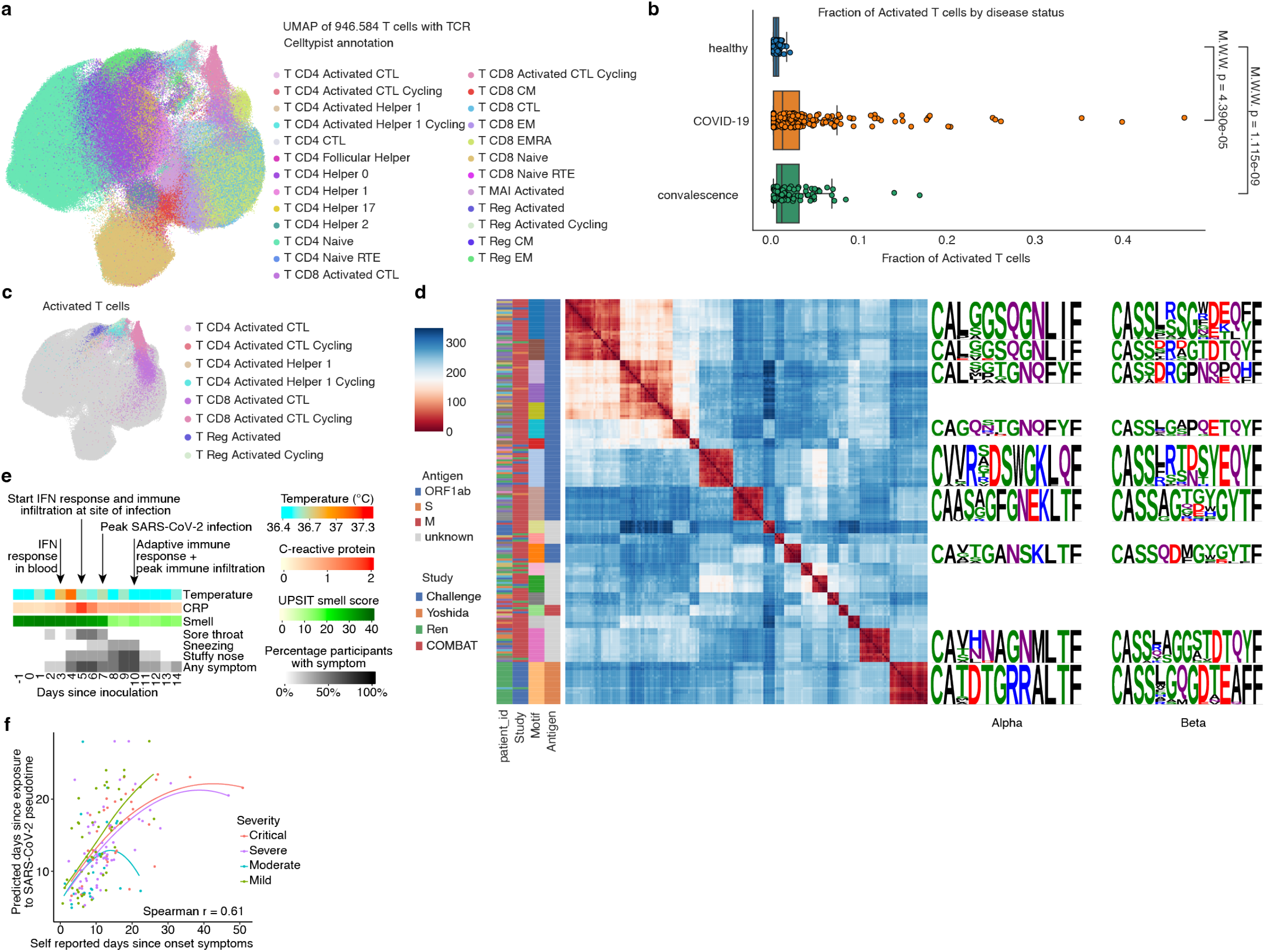
Integrating COVID-19 patient data reveals public SARS-CoV-2 TCR motifs. (**a**) UMAP representation after integration of five COVID-19 patient datasets with paired RNA and VDJ sequencing data. Cell type labels inferred using a logistic regression classifier (Celltypist) trained on manual annotations of PBMCs from the current work. (**b**) Fraction of activated T cells across all T cells in sample for COVID-19 (n = 240), convalescent (n = 82) and healthy (n = 88) samples of five COVID-19 patient datasets. Significance levels after Mann-Whitney testing are shown and indicate that COVID-19 and convalescent samples have significantly more activated T cells than healthy samples. (**c**) Activated T cell types highlighted on UMAP representation from panel (a). Activated CD8+ T cells were most abundant, followed by CD4+ and regulatory types, and clustered together in a distinct area of the latent space. (**d**) Clustermap of pairwise TCR distances with color-coded information for each TCR on patient_id, study, motif, antigen on the left-hand side, as well as the sequence logos for the nine most common motifs on the right-hand side. Each column/row corresponds to a unique TCR, and the distance to each TCR in the set is indicated by color. Only activated T cells with public motifs (identified in more than one individual) were considered. Low distances indicate similar TCRs, with distances of 40 and less potentially yielding TCRs recognising the same epitopes. For sequence logos, letter height indicates frequency of AA at that position across T cells pertaining to the motif. AAs are colored by side chain chemistry: Acidic (red), basic (blue), hydrophobic (black), neutral (purple), polar (green). AA: amino acid. (**e**) Recorded symptoms averaged over SARS-CoV-2 challenge participants with sustained infection for days -1 to 14 post-inoculation. Major molecular events in the immune response are highlighted with arrows. (**f**) Predicted time since viral exposure is plotted against reported time since onset of symptoms. Lines represent loess fits of the data split and color coded by reported severity.

We then employed our cell state aware clonotype group selection approach (Cell2TCR) to identify activated clonotypes, which resulted in 254 COVID-19 associated clonotype groups (**Extended Data Table 1f**). Strikingly, 211 of these activated clonotype groups were shared between patients (largest groups shown in **Fig. 4d**), highlighting the antigen-specificity of this approach, with the two most common motifs being shared by 13 individuals each. This also implies that a relatively small set of highly immunogenic SARS-CoV-2 peptides results in most of the T cell responses in COVID-19. Finally, we wanted to validate the antigen-specificity of the COVID-19 associated clonotype groups that we found in the public and challenge study data. Thus we intersected the CDR3 amino acid sequences with databases containing experimentally validated SARS-CoV-2 specific TCRs (see *Methods*). Importantly, this revealed that activated clonotype groups, including groups that contain TCRs from this study, are 3.75 fold enriched (p = 1.68 × 10^-21^) for SARS-CoV-2 specific TCRs. This provides strong validation for activated T cells indeed representing the antigen-specific T cell response against SARS-CoV-2 (**Extended Data Fig. 6i**). Most of the activated T cell clonotype groups recognise viral proteins encoded by ORF1ab, but we also find Membrane and Spike specific TCR clonotype groups. Because our cell state aware clonotype selection method identifies SARS-CoV-2 specific TCRs without any prior antigen information, our results may also include TCRs that recognise SARS-CoV-2 antigens that have not yet been tested. Together, these results validate the specificity of the adaptive immune response that we observed at day 10, and highlight the power of defining activated T cells for detecting disease-specific antigens in an unbiased manner.

### Molecular responses precede and are dynamic during clinical manifestations

Last, we wanted to investigate how our single-cell resolved timeline of immune responses relates to clinical manifestations that are typically observed and measured in COVID-19 patients. The unique experimental setting of our human challenge model enabled us to collect highly detailed and time-resolved clinical data for all participants that were profiled using single cell genomics approaches. The timing of the most relevant and dynamic COVID-19 symptoms show that even the earliest symptoms appear mostly at day 4 post-inoculation (**Fig. 4e**), which is later than some of the molecular responses that we described. Here, the upregulation of APR in ciliated cells, the activation of MAIT cells, depletion of some myeloid cells, and the global activation of IFN signaling in blood are observed before or at day 3 post challenge (**Fig. 2**). In contrast, a slight rise in temperature is only significantly detectable at day 4 post-inoculation (p = 5 × 10^-6^), at which early upper airway-related symptoms such as a stuffy nose and a sore throat also appear. This is then followed by global immune infiltration and activation of IFN signaling at the site of infection at day 5, which is also the first time that we detected infected cells. This coincides with a slight increase in C-reactive protein (CRP) in blood (p = 0.04). At day 7 post-inoculation we observed that the amount of detectable infected cells peaked. Interestingly, from day 8 on we also observed that all but one of the participants with a sustained infection significantly lost their sense of smell (p = 0.004), together with aggravation of sneezing and a stuffy nose. This was followed by a strong reduction in the amount of infected cells at day 10 and a peak in the amount of nasopharyngeal immune infiltration, which coincides with the onset and expansion of an adaptive immune response and clearance of most symptoms. In summary, we observe that clinical manifestations and different waves of immune responses dynamically change over time, which can aid the molecular interpretation of COVID-19 based on clinical observations and improves our understanding of the therapeutic time windows in this disease.

### Human COVID-19 challenge data as a reference atlas for cell dynamics

To maximize the impact of our time-resolved COVID-19 dataset, we build predictive models to infer time since SARS-CoV-2 exposure. We used Gaussian process regression and latent variable models to fit the changes in cell state composition during sustained infection. We next applied these predictions to publicly available PBMC single cell RNA-seq datasets from 361 COVID-19 samples, to infer at which stage of the immune response each patient was and to predict when this patient was exposed to SARS-CoV-2. Reassuringly, our Gaussian processes based time inference model predicts that the time since exposure and the time since onset of symptoms are highly correlated (**Fig. 4f**), and that exposure is predicted to precede onset of symptoms, as expected. Interestingly, the predicted difference between exposure and symptoms decreases with increased severity (**Extended Data Fig. 6j**), where patients with more severe COVID-19 are predicted to be in the adaptive immune reaction phase for longer. While this suggests that patients with severe COVID-19 take longer to clear the virus, it could also indicate that the cellular composition and immune response timeline in severe cases is perturbed compared to the relatively mild cases observed in the challenge study. In addition to a temporal model that could improve the assessment of the disease stage of COVID-19 patients, we also provide annotation models for a total of 202 cell states including new temporal and rare cell states. These models are now included in the default models at Celltypist.org, and enable highly detailed cell type annotation without the need for bioinformatics expertise. In addition, our single cell expression data is freely available at our COVID19CellAtlas.org web portal for online exploration and analysis.

## Discussion

Our findings have implications for the COVID-19 and broader infection diseases community. We detect multiple response states that precede the onset of clinical manifestations, including the activation of MAIT cells and decreases in inflammatory monocytes. Importantly, this represents a newly discovered immune response that emerges when an individual is exposed to SARS-CoV-2, but manages to prevent the onset of viral spread. These features of very early and abortive infections can be used as biomarkers and help to understand the immediate immune response upon viral exposure. In addition, we discovered a distinct cell state for activated conventional T cells that harbor SARS-CoV-2 specific TCRs, and we show that this signature can be projected onto patient cohort data to unravel the disease specific T cell response.

The timing of our challenge experiments in the early stages of a pandemic with a new virus - before most of the population acquired immune memory through natural infections and vaccine rollout - enabled us to recruit and study immune responses in adult participants that were completely naive to this pathogen. The resulting unique data will be essentially impossible to replicate in future efforts as the population builds memory to many SARS-CoV-2 strains. In addition to the responses during sustained infections and COVID-19, we were also able to study abortive and transient infections that would be extremely challenging to detect outside a controlled challenge setting, revealing novel immune response signatures associated with successfully preventing sustained infections.

Our results uncovered a temporally resolved timeline of antiviral responses in epithelial and immune cells both locally at the site of infection and systemically in circulation (**Fig. 5**). During sustained infections that lead to COVID-19, we observe an immediate and novel acute phase response in ciliated cells at the site of infection. In blood, we observe two novel innate immune responses in which MAIT cells become activated, and inflammatory monocytes migrate out of circulation. These two responses are the only immune responses that are also observed in participants with abortive infections that did not develop COVID-19, underscoring their importance and sensitivity, and uncovering a new and distinct immune response to SARS-CoV-2 exposure that does not lead to COVID-19.

**Figure 5:**
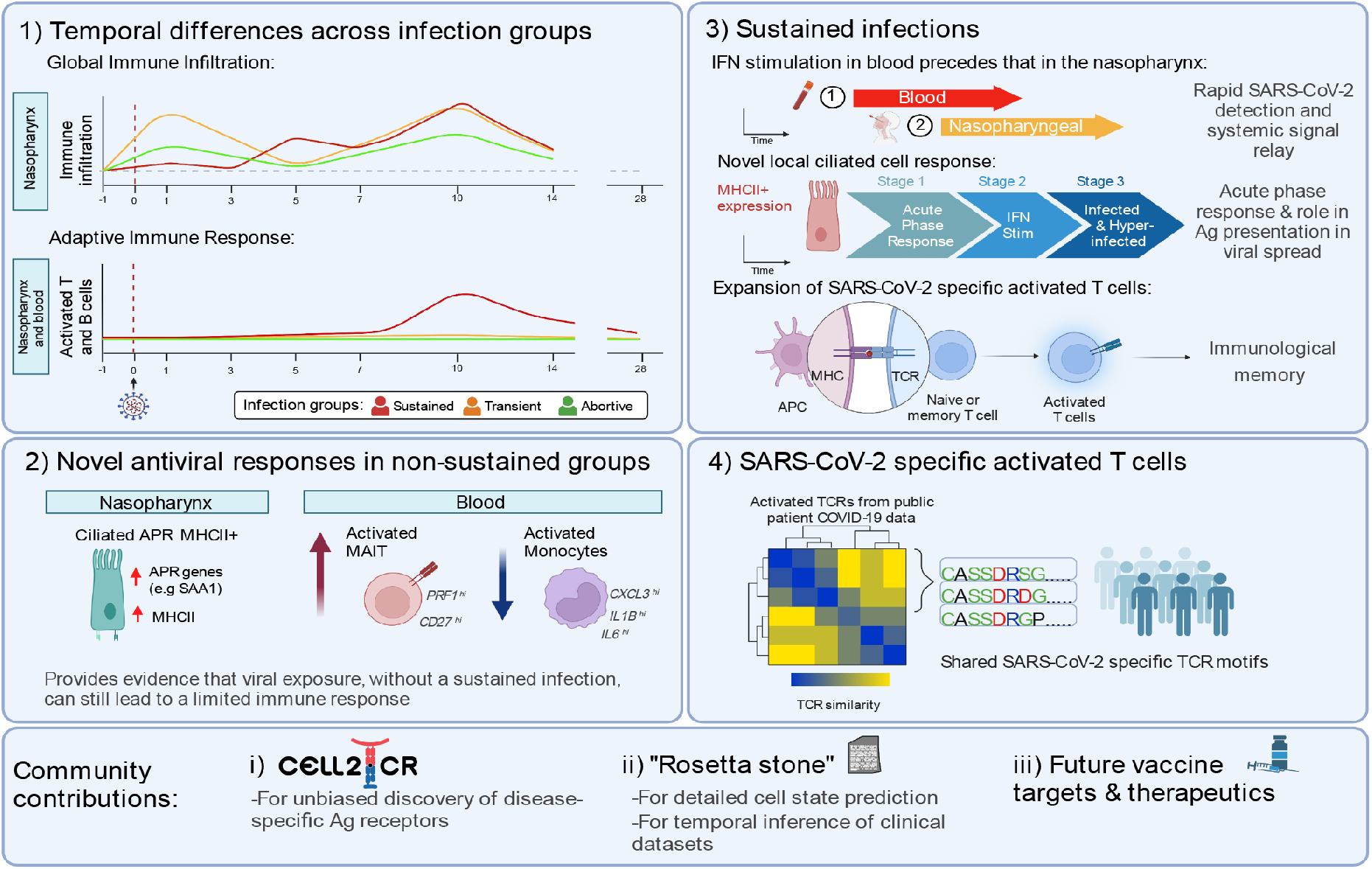
Temporally resolved epithelial and immune response in SARS-CoV-2 infections. Summary figure highlighting 1) temporal differences in the distinct infection groups, 2) novel antiviral responses, 3) novel characteristics of sustained infection, and 4) the identification of public motifs in SARS-CoV-2 specific activated T cells. In addition, our work provides community tools for inference of specific TCR motifs (Cell2TCR) in activated T cells, and for temporal assignments of clinical COVID-19 samples underpinning future therapeutic applications.

In sustained infections, we observe global activation of interferon signaling that affects all circulating immune cells. Strikingly, the activation of interferon signaling in blood precedes widespread activation at the site of inoculation, which suggests that a highly efficient relay to the systemic immune system exists, likely through the lymphatic system. The activation of interferon signaling at day 5 to seven post-inoculation coincides with global immune infiltration and a peak of detectable virally infected cells. This relatively slow immune response at the site of inoculation is in contrast to the immediate immune infiltration that we observed in infections that were only transiently detectable. In sustained infections, we also detect large amounts of cells containing viral RNA including infection of immune cells, but we provide evidence that only epithelial cells support successful viral replication. Here, we found that a small subset of hyper-infected ciliated cells becomes anti-inflammatory and the main source of viral production.

While our experimental approach included matched preinfection samples and we profiled all expected cell types from a total of 181 samples from 16 participants, we cannot exclude the possibility that our infection group sizes remained underpowered to detect very subtle or time-restricted responses. In addition, we note that the participants enrolled in this study cleared the infection with mild symptoms. It is possible that COVID-19 patients that require hospitalization exhibit perturbed or exacerbated immune responses that were not captured in our work, which could mean that caution should be taken when extrapolating our findings to critically ill COVID-19 patients.

At day 10 post-inoculation, we detect the onset and expansion of the adaptive immune response. In addition to antibody-secreting B cells, this response also includes activated conventional T cells. This is the first time to our knowledge that these cells have been described in single cell transcriptomics assays, likely because of the limited time window in which these activated T cells are detectable. Two weeks after inoculation, the amount of activated regulatory T cells at the site of inoculation peaks, while the abundance of other immune cells normalizes again, which coincides with a near absence of any remaining infected cells. These activation states have key marker genes, and we can identify these activated CD4+, CD8+, and regulatory T cell states using machine learning models. We integrate their prediction into a computational pipeline (Cell2TCR) which includes paired chain TCR motif inference. This is a tool applicable to any single cell RNA/VDJ dataset of infection, inflammation or tumor immune response.

Together, this represents the most comprehensive and detailed time-resolved description of the course of SARS-CoV-2 infection, or any other infectious disease, and gives unique insights into responses that are associated with resisting a sustained infection and disease.

## Data Availability

The data presented in this study can be explored and analyzed interactively through our COVID-19 Cell Atlas web portal (https://covid19cellatlas.org), currently accessible via early-access at http://covid19-challenge-study.cellgeni.sanger.ac.uk/ . The cell by feature count matrices are also available to download at the web portal. The cell state annotation models are available in the CellTypist model repository (https://www.celltypist.org/models). A reference for our MT-GPR model to infer time since viral exposure on PBMC data is available at our GitHub repository (https://github.com/Teichlab/COVID-19_Challenge_Study). The raw sequencing data will be made available via the European Genome-Phenome Archive, which will be made available under managed data access. Cell2TCR is available at our GitHub repository (https://github.com/Teichlab/Cell2TCR). Code that was used for data analysis is available at our GitHub repository (https://github.com/Teichlab/COVID-19_Challenge_Study).

## Acknowledgements

We are grateful to Sheila Casserlya and Sujana Regmi for assistance with collecting samples, and Tarryn Porter, Agnes Oszlanczi, Yvette Wood and Sabine Eckert for library preparation. We thank Rasa Elmentaite for helpful discussions on activated T cells. We acknowledge assistance from Rea Dabelić (10X Genomics), Illumina and 10X Genomics. We are grateful for the support and guidance with MACS for the Dextramer work provided by Yanping Guo, the Flow Cytometry Translational Technology Platform Manager at UCL Cancer Institute. We acknowledge assistance provided by the University College London CL3 facility at the Paul O’Gorman building and the staff at the Sanger Institute Core Sequencing facility.

This research was funded in whole, or in part, by the Wellcome Trust Grant 206194, 220540/Z/20/A and 211276/Z/18/Z, and by Action Medical Research (GN2911, to M.Z.N. and K.B.M.). M.Z.N. acknowledges funding from a MRC Clinician Scientist Fellowship (MR/W00111X/1). M.Z.N. and J.L.B. acknowledge funding from the Rutherford Fund Fellowship allocated by the MRC UK Regenerative Medicine Platform 2 (MR/5005579/1). M.Y. was funded by The Jikei University School of Medicine and Action Medical Research (GN2911). K.B.W. acknowledges funding from University College London, Birkbeck MRC Doctoral Training Programme. L.M.D is supported by the European Union’s Horizon 2020 research and innovation programme under the Marie Skłodowska-Curie grant agreement No 955321. M.N. acknowledges funding from the Wellcome Trust (207511/Z/17/Z) and by NIHR Biomedical Research Funding to UCL and UCLH. R.H. is a NIHR Senior Investigator.

For the purpose of Open Access, the author has applied a CC BY public copyright license to any Author Accepted Manuscript version arising from this submission.

## Author Contributions

M.Z.N. and S.A.T. conceived, set up, directed this study and provided funding. C.C. set up the clinical study and co-ordinated sampling. K.B.W. optimized digestion protocols, processed samples for 10X and CITEseq, isolated DNA for genotyping, performed Dextramer experiments and assisted with data analysis and interpretation. R.G.H.L. performed and led the data analysis. L.M.D. assisted with data analysis. R.G.H.L., K.B.W., L.M.D., K.B.M., M.Z.N. and S.A.T. interpreted the data and wrote the manuscript. M.Y., J.L.B., J.A.H. assisted with 10X sample processing. V.T., S.M.J. and R.C.C. provided student supervision to K.B.W. and PM.. H.R.W. processed blood samples. P.M. collected nasopharyngeal samples for optimisation of digestion protocols. M.H., M.N. and R.H. assisted in the set up of the study. B.K., M.K., A.C., A.B. and M.M. oversaw sample collection and provided clinical data.

These authors contributed equally: Rik G.H. Lindeboom, Kaylee B. Worlock

These authors jointly supervised this work: Marko Z. Nikolić, Sarah A. Teichmann

## Data Availability

The data presented in this study can be explored and analyzed interactively through our COVID-19 Cell Atlas web portal (https://covid19cellatlas.org), currently accessible via early-access at http://covid19-challenge-study.cellgeni.sanger.ac.uk/ until publication in a peer reviewed journal. The cell by feature count matrices are also available to download at the web portal. The cell state annotation models are available in the CellTypist model repository (https://www.celltypist.org/models). A reference for our MT-GPR model to infer time since viral exposure on PBMC data is available at our GitHub repository (https://github.com/Teichlab/COVID-19_Challenge_Study). The raw sequencing data will be made available before publication in a peer-reviewed journal via the European Genome-Phenome Archive, which will be made available under managed data access.

## Code availability

Cell2TCR is available at our GitHub repository (https://github.com/Teichlab/Cell2TCR). Code that was used for data analysis is available at our GitHub repository (https://github.com/Teichlab/COVID-19_Challenge_Study).

## Competing interest statement

R.G.H.L., L.M.D. and S.A.T. are inventors on a filed patent that is related to the detection and application of activated T cells. In the past three years, S.A.T. has received remuneration for Scientific Advisory Board Membership from Sanofi, GlaxoSmithKline, Foresite Labs and Qiagen. S.A.T. is a co-founder and holds equity in Transition Bio. P.M. is a Medical Research Council (MRC)-GlaxoSmithKline EMINENT clinical training fellow with project funding unrelated to the topic of this Comment and receives co-funding from the National Institute for Health Research (NIHR) University College London Hospitals (UCLH) Biomedical Research Centre. P.M. reports consultancy fees from SOBI, AbbVie, UCB, Lilly, Boehringer Ingelheim, and EUSA Pharma all unrelated to this submission. A.M., A.C., M.K., M.M. and A.B. are full time employees at hVIVO Services Ltd.

## Methods

### Study participants and design

16 healthy adults aged 18-30 years, with no evidence of a previous SARS-CoV-2 infections or vaccinations (seronegative), were included for this study from the wider cohort (34 participant) enrolled as part of the Human COVID-19 Challenge study, pioneered by the government task force, Imperial college london, Royal Free London NHS Foundation Trust and hVIVO^6^. These participants were enrolled as part of the cohort 5 and 6, from June - August 2021. Reported patient identifiers have been de-identified and are not known by the participants in this study. Volunteers were tested for the presence of anti-SARS-CoV-2 protein antibodies via the MosaiQ COVID-19 antibody microarray (Quotient) prior to enrollment and excluded based upon a positive test, as well as, upon risk factors assessed by clinical history, physical examinations and screening assessments. See Killingley, Mann et al, (2022)^6^ for the full list of inclusion and exclusion criteria and for further details regarding the challenge set up and ethics. In short, written informed consent was obtained from all volunteers before screening and study enrollment. The clinical study was registered with ClinicalTrials.gov (identifier NCT04865237). This study was conducted in accordance with the protocol, the Consensus ethical principles derived from international guidelines including the Declaration of Helsinki and Council for International Organizations of Medical Sciences (CIOMS) International Ethical Guidelines, applicable ICH Good Clinical Practice guidelines, applicable laws and regulations. The screening protocol and main study were approved by the UK Health Research Authority – Ad Hoc Specialist Ethics Committee (reference: 20/UK/2001 and 20/UK/0002).

Participant 674007, who fulfilled enrollment criteria, was later found to have low levels of neutralizing and spike-binding antibodies on admission to the quarantine upon more sensitive post-study experiments and analysis. This patient was classified as an abortive infection based on the virus kinetics (see virology method below), with the exclusion of this individual found not to alter any of our conclusions.

The participants were followed for 1 year after inoculation, with continued samples and metadata collected for the use in future/further studies and to benefit the research community. This study however focused primarily on the first 28-days post-inoculation (with the exception of 46 days for one participant as noted below, see Sample collection).

### Challenge virus

Participants were inoculated intranasally with an wild-type pre-alpha SARS-CoV-2 challenge virus (full formal name: SARS-CoV-2/human/GBR/484861/2020) at dose 10 TCID50 at day 0. 100 µl per naris was pipetted between both nostrils and the participant was asked to remain supine (face and torso facing up) for 10 minutes, followed by 20 minutes in a sitting position wearing a nose clip after inoculation to ensure maximum contact time with the nasal and pharyngeal mucosa. Mid-turbinate nose and throat samples were collected twice daily using flocked swabs and placed in 3 ml of viral transport medium (BSV-VTM-001, Bio-Serv) that was aliquoted and stored at −80 °C in order to evaluate viral kinetics (infection status) as described in Virology method section below. Participants remained in quarantine for a minimum of 14 days post-inoculation until the following discharge criteria were met: two consecutive daily nose and or throat swabs with no viral detection or a qPCR Ct value >33.5 and no viable virus by overnight incubation viral culture with detection by immunofluorescence. For protocol/full details and ethics used within the Human SARS-CoV-2 challenge study see the Challenge virus section of methods in Killingley and Mann., et al (2022)^6^.

### Sample collection

#### Nasopharyngeal swabs

Samples were collected at the Royal Free Hospital by trained healthcare providers at 7 timepoints; day-1 (pre-inoculation) and day 1, 3, 5, 7, 10 and 14 post-inoculation. The patients were asked to clear any mucus from their nasal cavities and nasopharyngeal samples were collected using FLOQSwabs (Copan flocked swabs, Ref 501CS01) inserted along the nasal septum, above the floor of the nasal passage to the nasopharynx until a slight resistance was felt. The swab was then rotated in this potion in both directions for 10 seconds and slowly removed whilst still rotating and immediately stored in a pre-cooled cryovial on wet ice containing freeze media (90% heat inactivated fetal bovine serum (FBS) and 10% dimethyl sulfoxide (DMSO). On wet ice the cryovials vials were the transferred to the hospital chutes where they were sent down to the laboratory (<2 mins at RT) and placed in a slow-cooling device (Mr. Frosty Freezing Container, Thermo Fisher Scientific) and stored at −20 °C until all samples were collected, at which point they were moved to −80 °C freezers for at least 48 hours for optimum freezing. Samples were moved, stored and in liquid nitrogen for later processing.

#### PBMC isolation from peripheral blood

Peripheral whole blood was collected at the Royal free hospital in EDTA tubes at 5 timepoints; day-1 (pre-inoculation) and day 3, 5, 10, 14 and 28 post-inoculation. Each day the blood was transferred at room temperature to Imperial College London for the fresh isolation and collection of peripheral blood mononuclear cells (PBMC) by means of Histopaque Ficoll separation (Merck, H8889-500ML). The peripheral whole blood was first diluted 1:1 with 1X PBS (Merck, D8662-500ML) before being gently overlayed onto a maximum of 15 mL of Histopaque, at a ratio of 2:1 (blood:Histopaque). The samples were then centrifuged at 400x*g* (with no breaks) for 30 min at room temperature (RT) and the PBMC white buffer layer collected, washed (with PBS ∼50 mL) and spun down (400x*g* for 10 min at RT), before the supernatant was carefully discarded and the cell pellet was resuspended in 10 mL PBS. The cells were filtered using a 40 or 70mm cell strainer and then both the cell number and viability were assessed using Trypan Blue. The cells were further centrifuged (400x*g* for 10 mins) and resuspend in the required volume of Cell Freezing Media; 90% FBS (Sigma, F9665-500ML), 10% DMSO (Sigma, D2650-100ML), before being cryopreserved at −80 °C using a slow-cooling device. The blood and nasopharyngeal samples were collected within 2 hours of each other.

Of note after their were discharged from quarantine and prior to their 28-day follow up (where additional blood samples were collected), two participants reported either to have had their first SARS-CoV-2 vaccine (636163) or a community infection; 636163 had their first vaccine on day 14 post-inoculation (2 weeks before the day-28 sample was taken). 677306 tested positive before their day 28 visit was due. The follow up was therefore delayed by 2 weeks, resulting in the “day 28” sample for this participant instead being taken day 46 post-inoculation. ELISpot performed on this participant revealed a response in the day 28 and 90 samples (data not shown). Moreover, participant 677696 tested positive on day 29 post-inoculation, a day after their day 28 sample was taken. However for this participant the ELISpot showed no response at day 28 and a small response at day 90 suggesting the day 28. See **Extended Data Fig. 1a** for overview of the samples and timepoints included from each participant. These individuals/timepoints were found not to alter any of our conclusions.

### Clinical assessments

Participants were carefully monitored and assessed daily using an array of blood tests, spirometry, electrocardiograms and clinical assessments (vital signs, symptom diaries and clinical examination). Full details of all the safety and clinical data collected with the human SARS-CoV-2 challenge study can be seen in the methods in Killingley and Mann., et al (2022)^6^, with an overview of select metadata for the 16 participants enrolled in this study in **Extended Data Table 1g**.

### Virology

Longitudinal measures from the nose and throat (pharyngeal) were carried out daily in order to assess and quantify the viral kinetics of each participant pre- and post-inoculation. These were measured using two independent assays: (1) qRT–PCR with N gene primers/probes adapted from the Centers for Disease Control and Prevention (CDC) protocol^27^ (updated 29 May 2020) and (2) quantitative culture by Focus Forming Assay (FFA). For full details of each assay and statistical analysis refer to the methods in Killingley and Mann., et al (2022)^6^.

The lower limit of quantification (LLOQ) for RT-qPCR was 3 log10 copies per milliliter, with positive detections less than the LLOQ assigned a value of 1.5 log10 copies per milliliter and undetectable samples assigned a value of 0 log10 copies copies per milliliter. Only samples where participants presented with a positive RT-qPCR were further tested using the FFA assay. In the FFA the LLOQ was 1.27 FFU ml−1; viral detection less than the LLOQ was assigned 1 log10 FFU ml−1; and undetectable samples were assigned 0 log10 FFU ml−.

A sustained laboratory-confirmed infection was defined as quantifiable RT–qPCR detection greater than the LLOQ from mid-turbinate and/or throat (pharyngeal) swabs on two or more consecutive 12-hourly time points, starting from 24 hours after inoculation and up to discharge from quarantine. Participants where only stand alone RT–qPCR tests returned quantifiable results (**>** LLOQ) were classified as transient infections. Participants where no RT–qPCR tests returned quantifiable results (**>** LLOQ) were classified as abortive infections (See **Extended Data Fig. 1b** and **Extended Data Table 1a,b,h,i**).

Infection intervals for each participant were calculated based on the time of the first and last RT-qPCR test with detectable virus (across the nose and/or throat), where timepoints in which tests below the LLOQ (1.5) were also counted if they occurred < 2 days of a quantifiable (> LLOQ) test result.

### Nasopharyngeal swab dissociation and processing

Following freezing, nasopharyngeal swabs were transferred to a category level 3 facility at University College London were stored and processed in batches of 7-to -8 samples at a time to a single cell suspension. All work was carried out in a MSC class I hood in compliance with standard category level 3 safety practices. The dissociation and collection of cells from nasopharyngeal swab was carried out in accordance with the previously described protocol^28, 29^, with minor modifications. This approach involves multiple parallel washes and digestion steps using both the nasopharyngeal swab and collected freezing/ wash media to help ensure the maximum cells and cellular material is collected. First samples are exposed to DTT for 15 mins, followed by an accutase digestion step for 30 mins before cells from the same sample (collected directly from the swab or the freezing media/washes from that swab) are quenched, pooled and filtered prior to checking the cell number and viability.

Briefly, samples were rapidly thawed and the liquid collected in an empty 15mL falcon tube (**Tube B**). The cryovial, lid and swab was then carefully rinsed with 3x 1mL warm RPMI 1640 medium which was added dropwise to the 15 mL tube whilst gently swirling the tube, in order to slowly dilute the DMSO from the freezing media to help present the cells bursting. After waiting 1 min, the tube (**Tube B**) was then topped up with an extra 2 mL of warm RPMI 1640 media and centrifuged at 400 g for 5 min at 4°C. The cell pellet was then resuspended in RPMI 1640/10 mM DTT (Thermofisher, R0861), and incubated for 15 min on a thermomixer (37°C, 700 rpm), centrifuged as above and the supernatant was aspirated and the cell pellet was resuspended in 1 mL Accutase (Merck, A6964-500ML). This was then incubated for a further 30 min on the thermomixer (37°C, 700 rpm).

In parallel to the processing of the cell freezing media/washes above, the swab was moved to an new 1.5 mL eppendorf tube (**Tube C**) containing 1 mL RPMI 1640/10 mM DTT and placed on the thermomixer (37°C, 700 rpm) for 15 minutes. In accordance with the steps above, the swab was next transferred to a new 1.5 mL eppendorf (**Tube D**) containing 1 mL Accutase and incubated with agitation (700 rpm) at 37°C. The 1 mL RPMI 1640/10 mM DTT from the nasopharyngeal swab incubation (in **Tube C**) was centrifuged at 400 g for 5 min at 4°C to pellet cells, the supernatant was discarded, and the cell pellet was resuspended in 1 mL Accutase and incubated for 30 min at 37°C with agitation (700 rpm).

Following the Accutase digestion step, all cells were combined (Tube B, C and D) and filtered using a 70 µm nylon strainer (pre-wetted with 3 mL quenching media: RPMI 1640/10% FBS/1 mM EDTA (Invitrogen, 1555785-038) in a 50 mL conical tube (**Tube E**).

The filter, tubes and swab were then further thoroughly rinsed with quenching media in order to collect all cells and the washes combined. The dissociated, filtered cells (Tube E) were then centrifuged at 400 g for 5 min at 4°C, and supernatant discarded. The cell pellet was resuspended in residual volume (∼500 µL) and transferred to a new 1.5 mL eppendorf tube (**Tube F). Tube E** was then washed with a further 500 µL of RPMI 1640/10% FBS and combined with Tube F, centrifuged as above, supernatant removed and cells resuspended in 20 µL of RPMI 1640/10% FBS. Using Trypan Blue, total cell counts and viability were assessed. The cell concentration was adjusted for 7,000 targeted cell recovery according to the 10x Chromium manual before loading onto the 10x chip (between 700–1,000 cells per µl) and processing immediately for 10x 5′ single-cell capture using the Chromium Next GEM Single Cell V(D)J Reagent Kit v1.1 (Rev E Guide). For samples where fewer than 13,200 total cells were recovered, all cells were loaded.

Note: Due to the sample type, necessary freezing process and no access to a class 3 flow facility to sort out viable cells, the majority of the samples processed were seen to have low viability (ranging from 5.4% viability to 57.85, with the average viability of samples processed of 26.89%).

### PBMC CITE-seq staining for single-cell proteogenomics

Frozen PBMC samples were thawed and processed in batches of 16 to allow for a carefully designed pooling strategy. Here each sample was pooled twice into two unique pools containing up to four PBMC samples per pool from mixed timepoints. Note: Only one sample from each donor was even pooled together at a time to assist with the demultiplexing later. This pooling strategy was used to help remove and correct for any protocol-based batch effects.

In brief, PBMC samples were thawed quickly at 37 °C in a water bath. Warm RPMI 1640 medium (20– 30 mL) containing 10% FBS (RPMI 1640/FBS) was added slowly to the cells before centrifuging at 300*g* for 5 min. This was followed by a wash in 5 mL RPMI 1640/FBS. The PBMC pellet was collected, and the cell number and viability were determined using Trypan Blue.

PBMCs from 4 different donors were then pooled together (1.25 × 105 PBMCs from each donor) to make up 5.0 × 105 cells in total. The remaining cells were used for DNA extraction (Qiagen, 69504). The pooled PBMCs were resuspended in 22.5 µl of cell staining buffer (BioLegend, 420201) and blocked by incubation for 10 min on ice with 2.5 µl Human TruStain FcX block (BioLegend, 422301). The PBMC pool was then stained with TotalSeq-C Human Cocktail, V1.0 antibodies (BioLegend, 399905) according to the manufacturer’s instructions (1 vial per pool). For a full list of TotalSeq-C antibodies (130 abs + 7 isotype controls) refer to **Extended Data Table 1j**. Following a 30 min incubation period with the TotalSeq-C Human Cocktail V1.0 antibodies (at 4 °C in the dark) the PBMCs were topped up using cell staining buffer and centrifuged down to a pellet (500*g* for 5 min at 4 °C) and discarding the supernatant. The pellet was then resuspended and washed in the same manner two more times using the resuspension buffer (0.05% BSA in HBSS), before finally being resuspended in a 20-30 µL resuspension buffer and counted again. The PBMC pools were then processed immediately for 10x 5′ single cell capture (Chromium Next GEM Single Cell V(D)J Reagent Kit v1.1 with Feature Barcoding technology for cell Surface Protein-Rev D protocol). 25,000 cells were loaded from each pool onto a 10x chip.

### PBMC Dextramer staining for SARS-CoV-2 antigen specific T cell enrichment and single-cell sequencing

In order to further validate and investigate the SARS-CoV-2 antigen-specific T-cell populations in our single cell dataset Day 10, 14 and 28 post-inoculation PBMCs samples, from all 16 participants, were further enriched and processed for single cell sequencing using a multi-allele panel of 44 SARS-CoV-2 antigen specific dCODE™ Dextramer® (10x compatible) (Immudex, see **Extended Data Table 1k** for full panel). This panel includes; five antigen-specific T-cell populations, spanning four MHCI and one MHCII alleles (covering a total of 15 participants, see **Extended Data Table 1l**) and several negative controls. Samples were then stained with several FACS antibodies (for monocyte and T cells) and sorted using MACSQuant® Tyto® Cell Sorter (Miltenyi Biotec), where PE-dCODE Dextramer®-positive cells were collected and processed for 10x 5′ single cell capture. This allowed for the quantification of paired clonal TCR sequence and TCR specificity by overlaying single-cell V(D)J expression onto dCODE Dextramer®-positive cell clusters.

The dextramer staining protocol was taken from Immundex and optimised/adapted to suit our samples and pooling/staining strategy. In brief, the PBMC samples were thawed in batches of 7 to 8 samples and the cell number and viability for each sample calculated using Trypan Blue as previously described above. All cells from each sample were then pooled together in a fresh 1.5 mL eppendorf tube. Note: The pooling strategy here was such that only one sample per participant/donor was used per pool in order to enable de-multiplexing by genotype later on and each pool containing a mixture of timepoints to help reduce batch effect. In order to ensure the collection of as many cells as possible, each of the original sample tubes was then washed with 200 µl of staining buffer (1X PBS pH 7.4 containing 5% Heat inactivated FBS (Thermo Fisher Scientific, 10500064) and 0.1g/l Herring sperm DNA (Thermo Fisher Scientific, 15634017) and added to the pool. The tube was then topped up to 1.4 mLs with staining buffer and centrifuged down to a pellet (400*g* for 5 min at 4 °C). The supernatant was carefully removed and the cell pellet gently resuspended in a total of 30-40 µl staining buffer depending on pellet’s size, ready for staining.

In parallel the dCODE™ Dextramer® master mix was prepared (in the dark) as per manufacturer protocol. To help avoid aggregates, each individual dextramer reagent was first microcentrifuge at full speed for 5 mins before adding 2 µL from each dCODE™ Dextramer® specificity to a low-bind nucleus free 1.5 eppendorf tube (Eppendorf, 30108051) containing 8.8 µl 100µM d-Biotin (Avidity science, BIO200) (0.2 ul d-Biotin per number of dCODE™ Dextramer® specificity i.e. 44).The dCODE™ Dextramer® master mix was mixed by gently pipetting before the total volume (96.8 µl) was added to the resuspended cells. The sample was then thoroughly mixed and incubated at room temperature for 30 mins in the dark. Following the addition of anti-human CD14-FITC (Biolegend, 325603) and CD3-APC (Biolegend, 300458) (at 1:50) the cells were incubated for a further 20 mins (at room temperature in the dark) before being topped up to 1.4 mL with wash buffer (1X PBS pH 7.4 containing 5 % heat inactivated FBS). The cells were centrifuged down to a pellet (400*g* for 5 min at 4 °C) and the supernatant discarded. The wash step was then repeated X2, with the latter using the addition of 1.4 mL wash buffer + 1:5000 DAPI (Sigma) as live/dead stain. The supernatant was removed and the cell pellet resuspended in 4 mL FACS buffer ; 1X PBS, 1% FBS, 25mM HEPES (Thermo Fisher Scientific, 15630-056) and 1 mM EDTA. The samples were then filtered (35 µm nylon mesh cell strainer) and PE dCODE Dextramer®-positive cells sorted using a MACSQuant® Tyto® Cell Sorter as per manufacturer guide (Settings; Mix speed= 800 rpm, Chamber temperature= 4 °C, Pressure= 150hPA, Noise Threshold =14.40, Trigger Threshold= off). Note: In order to collect as many cells as possible during sorting the entire sample was run on the MACSQuant Tyto, with the negative run through collected and re-run a second time to ensure no true positives were lost. See **Extended Data Fig. 8a** for gating strategy for sorting. The PE dCODE Dextramer®-positive cells were then collected, centrifuged (400*g* for 5 min at 4 °C) and resuspended in resuspension media before counting the cells. The entire sample was then processed for 10x 5′ single cell capture (Chromium Next GEM Single Cell V(D)J Reagent Kit v1.1 with Feature Barcoding technology for cell Surface Protein-Rev D protocol). Where over 25,000 cells were collected the sample was split equally and loaded over two lanes.

In order to provide additional controls, participants with non-compatible HLA types, including one volunteer (674700) matching none of the HLA types for the multi-allele dCODE Dextramer panel, were also processed and used to determine background noise.

### Library generation and sequencing

The Chromium Next GEM Single Cell 5′ V(D)J Reagent Kit (V1.1 chemistry) was used for single-cell RNA-seq library construction for all nasopharyngeal swab samples, and the Chromium Next GEM Single Cell V(D)J Reagent Kit v1.1 with Feature Barcoding technology for cell surface proteins was used for PBMCs, both to process the PBMCs stained with CITE-sequencing antibodies panel and the dCODE™ Dextramer® (10x compatible) panel. GEX and V(D)J libraries were prepared according to the manufacturer’s protocol (10x Genomics) using individual Chromium i7 Sample Indices. Additional TCR γ/δ enriched libraries were also generated based upon an in-house protocol previously described in ^30^. The cell surface protein libraries were created according to the manufacturer’s protocol with slight modifications used for the creation of libraries generated from the CITE-sequencing antibody panel. These included doubling the SI primer amount per reaction and reducing the number of amplification cycles to 7 during the index PCR to avoid the daisy chains effect. GEX, V(D)J and the CITE-sequencing derived cell surface protein indexed libraries were pooled at a ratio of 1:0.1:0.4 and sequenced on a NovaSeq 6000 S4 Flowcell (paired-end, 150 bp reads) aiming for a minimum of 50,000 paired-end reads per cell for GEX libraries and 5,000 paired-end reads per cell for V(D)J and cell surface protein libraries. The dextramer derived cell surface protein indexed libraries were submitted at a ratio of 0.1.

### Single cell genomics data alignment

Single-cell RNA-seq and CITE-seq data from PBMCs was jointly aligned against the GRCh38 reference that 10X Genomics provided with CellRanger 3.0.0, and alignment was performed using CellRanger 4.0.0. CITE-seq antibody-derived tag (ADT) barcodes were aligned against a barcode reference provided by the supplier, which we annotated to add informative protein names and made available in our GitHub repository. Single-cell RNA-seq data from nasopharyngeal swab samples were aligned against the same reference using STARSolo 2.7.3a, and post-processed with an implementation of emptydrops extracted from CellRanger 3.0.2. To detect viral RNA in infected cells, we added 21 viral genomes including pre-Alpha SARS-CoV-2 (NC_045512.2) to the above mentioned reference genomes for RNA-seq alignment, as described in Yoshida et al ^5^. Single cell alpha/betaTCR and BCR data was aligned using CellRanger 4.0.0 with the accompanying GRCh38 VDJ reference that 10X Genomics provided. Single cell gamma/delta TCR data was aligned against the GRCh38 reference that 10X Genomics provided with CellRanger 5.0.0, using CellRanger 6.1.2.

### Single cell genomics data processing

Both single cell RNA-seq and ADT-seq data were corrected using SoupX ^31^ to remove free-floating and background RNAs and ADTs. To correct ADT counts, SoupX 1.5.2 parameters soupQuantile and tfidfMin parameters were set to 0.25 and 0.2, respectively, and lowered by decrements of 0.05 until the contamination fraction was calculated using the autoEstCont function. SoupX on RNA data was performed using default settings. To confidently annotate SARS-CoV-2 infected cells, we used SoupX corrected viral RNA counts to remove false positives due to freely floating SARS-CoV-2 virions. However, when quantifying the amount of reads per cell in **Fig. 2h** and their distribution over the viral genome in **Fig. 2f**, we used the raw counts and sequencing data. To profile the distribution of viral reads, we removed PCR duplicates from the aligned BAM files that STARSolo produced with MarkDuplicates in picard (https://broadinstitute.github.io/picard/), and tallied the location within the SARS-CoV-2 genome using the start of each sequencing read. Aligned single cell RNA-seq data was imported from the *filtered_feature_bc_matrix* folder into Seurat V4.1.0 for processing, keeping only cells with at least 200 RNA features detected. Nasopharyngeal and PBMC cells with more than 50% and 10% of the counts coming from mitochondrial genes were excluded, respectively. SoupX corrected gene expression and ADT counts were normalized by dividing it by the total counts per cell and multiplying by 10 000, followed by adding one and a natural-log transformation (log1p).

### Demultiplexing and patient id assignment

Each PBMC sample was pooled twice into two unique pools containing up to four PBMC samples per pool, followed by CITE-seq and single cell VDJ sequencing as described above. Souporcell V2.0 ^32^ was used to demultiplex each pools based on the genotype differences between the mixed samples. Souporcell analyses were performed with the skip_remap parameter enabled and using the common SNP database that was provided by the software. We used two complementary approaches to confidently assign participant identity to each souporcell cluster. First we compared the cluster genotypes with SNP array derived genotyping data, generated for all participants and performed using the Affymetrix UK Biobank AxiomTM Array kit by Cambridge Genomic Services (CGS). Second, the combinations of samples within each pool was unique, enabling assignment of participant identity based on the presence of unique participant-specific combinations of identical genotypes in two separate pools. This multiplexing and replication strategy furthermore enabled us to distinguish library specific batch effects from participant specific effects in downstream analyses.

### Doublet detection

We used the output from souporcell to identify ground-truth doublets in PBMCs by selecting droplets that contained two genotypes from different participants. We then included these ground-truth doublets into the iterative rounds of subclustering and cell state annotation to look for doublet specific clusters that emerged, which we then subsequently removed. Doublets in the nasopharyngeal data were removed during iterative rounds of subclustering and cell state annotation by identifying cell clusters that expressed marker genes from multiple distinct cell types.

### Clustering and cell type annotation

Principal component analysis was run on corrected gene expression counts from selected hypervariable genes and the first 30 principal components were selected to construct a nearest neighbour graph and UMAP embedding. We used harmony^33^ to perform batch correction on the PBMC data on the sequencing library identity to remove technical batch effects. Leiden clustering^34^ performed at resolutions of 0.5, 1, 4 and 32, on nearest neighbour graphs and embeddings created with 500, 1000, 2000, 4000, 6000 and 8000 selected hypervariable genes (excluding TCR and BCR genes), was used to perform iterative rounds of cell type annotation based on marker gene expression and subsetting of clusters to obtain a highly granular cell state annotation. We used cell type marker genes described in Yoshida et al^5^ and Stephenson et al^4^ to define cell types. Our cell type annotation was furthermore guided by predicted cell type labels using Celltypist’s^35^ provided models and custom trained models based on annotations in Yoshida et al^5^ and Stephenson et al^4^.

### Single cell TCR and BCR data processing

Aligned single cell BCR and alpha/beta TCR sequencing data was imported in scirpy^36^ to obtain a cell by TCR or BCR formatted table, which was then added to Seurat objects containing gene expression data. Aligned single cell gamma/delta TCR data was reannotated using Dandelion V0.2.4^37^.

### Integration of five COVID-19 studies

All transcriptomic data was processed with the single cell analysis Python workflow Scanpy^38^. Each data set was individually filtered following best practices outlined in^39^ (Between 200 and 3500 genes per cell, less than 10% mitochondrial genes expressed per cell, genes expressed in at least 3 cells, other parameters at default). The gene sets were reduced to their intersection before combining data sets. Cells came from a total of 602 individuals, with 325 acute COVID-19 patients, 110 COVID-19 convalescent patients, 114 healthy and 53 hospitalized controls (**Extended Data Table 1d**). This resulted in an integrated embedding containing 946,584 T cells with resolved TCR from 494 samples, made up of 455 donors of which 240 were acute COVID-19 patients, 82 convalescent, 88 healthy and 45 hospitalized controls (**Extended Data Table 1e**). The total number of donors in the integrated object is smaller as only samples with matching VDJ sequencing data were kept. A probabilistic scVI model (2 hidden layers, 128 hidden nodes, 20-dimensional latent space, negative binomial gene likelihood, other parameters at default^40^) was trained on the data to map cells to a shared latent space, and visualised using UMAP.

### Identification of activated TCR clonotype groups using Cell2TCR

To identify TCR clonotype groups, we used tcrdist3^41^ with the provided human references to compute a sparse representation of the distance matrices for all identified TRA and TRB CDR3 sequences, with the radius parameter set to 150. We then summed the distances for TRA and TRB to obtain a combined distance matrix. Next, we iterated over possible TCR distance thresholds between 5 and 150 with increments of 5, to compute TCR clonotype groups at each threshold. We then generated a distance adjacency graph of TCRs from different T cells with a distance lower than the threshold, which was clustered to identify TCR clonotype groups using leiden^34^ clustering through the igraph package^42^, at a resolution of 1 and using the RBConfigurationVertexPartition partition. To find the optimal distance threshold at which only TCRs that recognise the same antigen are grouped together, we quantified clonotype group contamination at each threshold using two approaches. First, we assumed that T cells that were annotated as naive should not participate in an expanded clonotype group, and quantified the proportion of naive T cells in each clonotype group to determine the largest threshold at which we observed minimal participation of naive T cells. Second, we assumed that CD4+ T cells and CD8+ T cells should never be part of the same TCR clonotype group, so we set out to quantify the proportion of CD4+/CD8+ mixing in each clonotype group to find the largest threshold where mixing is minimal. Both approaches revealed the same optimal threshold of 35 at which both naive T cell participation and CD4+/CD8+ mixing is minimal, which we then used for downstream analyses. To identify activated TCR clonotype groups we assumed that these groups should include activated T cells and that we should at least detect multiple independent TCR clonotypes that appear to be raised against the same antigen at the same time; we therefore selected clonotype groups that contained at least one participating activated T cell and that contained at least two unique CDR3 nucleotide sequences.

### Identification of activated BCR clonotype groups

To identify BCR clonotypes groups that were activated during infection, we used a similar approach as described above for T cells. Instead of using tcrdist to compute distances, we used the levenshtein distance and iterated over possible thresholds between 1 and 20 to find an optimal threshold by quantifying naive B cell participation. This revealed that a levenshtein distance of 2 is optimal to identify BCR clonotype groups that only contain B cells that recognise the same antigen. To identify activated BCR clonotype groups, we assumed that these groups should include antibody secreting B cells (plasmablasts and plasma cells) and that we should at least detect multiple independent BCRs clonotypes that appear to be raised against the same antigen at the same time; we therefore selected clonotype groups that contained at least one participating antibody secreting B cell and that contained at least three unique CDR3 nucleotide sequences.

### Generation of VDJ logos

TCR and BCR logos in Figure X, X and X were generated by providing the CDR3 amino acid sequences of each clonotype group to the ggseqlogo R package^43^ or the logomaker Python package^44^. When clonotype groups contained CDR3 amino acid sequences of variable lengths, we selected the sequences with the most frequently occurring length within each group for visualization purposes only.

### Generalized linear mixed models of cell state compositional changes over time

The relative amount of cells per cell type in each sample was modeled using a generalized linear mixed model with a poisson outcome. When technical replicates were available (most of the PBMC samples), these were modeled as separate samples. We modeled participant identifiers, days since inoculation, and sequencing library identifiers (of multiplexed libraries), as random effects terms to overcome colinearity between these factors. The effect of each clinical/technical factor on cell type composition was estimated by the interaction term with the cell type. The glmer function in the lme4 package implemented on R was used to fit the model. The standard error of the variance parameter for each factor was estimated using the numDeriv package. The conditional distribution of the fold change estimate of a level of each factor was obtained using the ranef function in the lme4 package. The log-transformed fold change is relative to the pre-inoculation time point (day -1). The statistical significance of the fold change estimate was measured by the local true sign rate, which is the probability that the estimated direction of the effect is true, that is, the probability that the true log-transformed fold change is greater than 0 if the estimated mean is positive (or less than 0 if the estimated mean is negative). We calculated P values using a two-sample Z-test using the estimated mean and standard deviation of the distribution of the effect (log-transformed fold change). P values were converted into false discovery rates using the Benjamini & Hochberg method.

### Gaussian processes regression and latent variable models to infer time since viral exposure

To infer time from cell state abundances, we first generated a linear regression model using CellTypist to predict PBMC or nasopharyngeal cell states based on the highly detailed manual cell state annotation presented in this work. Celltypist models were trained and used using default parameters, with check_expression set to false, balance_cell_type set to true, feature_selection set to true, and max_iter set to 150. We next set out to build a predictive model to infer time since viral exposure using the PBMC data presented in this work as a training dataset. We used the above mentioned publicly available PBMC data from five studies as a test dataset to predict time since viral exposure on. Because we were specifically interested in comparing time since viral exposure to reported time since onset of symptoms in varying disease severities, we excluded samples for which these features were unknown. To ensure that the cell state proportions in the training and test dataset were similar, we used our CellTypist model on both datasets to predict relative cell state frequencies, which was used as input for our time prediction model. To account for participant-to-participant heterogeneity and continuous variation in the timeline of immune responses, we first constructed a gaussian process latent variable model ^45^ (GPLVM) to smooth the time since viral exposure in the training dataset. We applied the Pyro implementation of GPLVM ^46^ across all predicted cell state abundances, and restricted the model to 2000 iterations and a single latent variable that was initialized on the square root transformed time since inoculation, which resulted in an accurate recapitulation of the mean time since inoculation while smoothing outliers. We next used each predicted cell state as input for a task to generate a multi-task gaussian process regression model^47^ to predict the smoothened time since inoculation using GPyTorch ^48^. We used the Adam optimiser and allowed for as many iterations for the loss in marginal log likelihood to reach zero. We next predicted the cell state compositions across the entire tested timeline (day -1 to day 28), and compared these cell state compositions to the cell state compositions in our query dataset as predicted by our CellTypist model. Last, we selected the time point whose predicted cell state composition had the lowest mean squared error compared to the observed cell state composition.

### Matching clonotype groups to antigen-TCR database

Computation of fold change enrichment of SARS-CoV-2 specific TCRs in Activated T cell populations compared to other T cell populations: Median p for 10 random draws of n=5000 unique clones of both populations = 3.75, p = 1.68 × 10^-21^

### Bulk TCR sequencing and processing

Total RNA was extracted from whole blood samples collected in Tempus Blood RNA tubes (Thermofisher #4342792) using the manufacturer’s protocol for RNA extraction. TCR α and β genes were sequenced using a pipeline which introduces unique molecular identifiers attached to individual cDNA molecules using single-stranded DNA ligation. The unique molecular identifier allows correction for sequencing error PCR bias, and provides a quantitative and reproducible method of repertoire analysis. Full details for both the experimental TCRseq library preparation (^49, 50^) and the subsequent TCR annotation (V, J and CDR3 annotation) using Decombinator V4^51^ are published. The Decombinator software is freely available at https://github.com/innate2adaptive/Decombinator.

### Memory formation analysis

T cell phenotypes (naive, activated, effector, memory) were recorded for an antigen-specific TCR clone at different time points throughout infection. TCR clones were filtered by having an activated label at least once, being observed in at least two samples, one of which had to be at day 28. Unique TCR clones are distinguished by color and numbered with their clone_id identifier. A shaded area is drawn when the same clone appeared with several distinct cell type labels, and the size of the shaded area informs of their relative ratios.

### Quantifying TCR diversity restriction in phenotypic clusters using coincidence analysis

To quantify the diversity of TCRs found within different phenotypic clusters we determined the probability with which two distinct clonotypes within a cluster share an identical CDR3 amino acid sequence ^52^. For visualization we normalized these probabilities by the same quantity calculated over the complete data regardless of phenotype. This ratio of probability of coincidences provides a stringent measure of convergent functional selection of distinct clonotypes that share the same TCR. The analysis is based on clonotypes defined by distinct nucleotide sequences of the hypervariable regions, and does not make direct use of clonal abundances as these can also reflect TCR-independent lineage differences. We focused our analysis on conventional T cells only, considered only cells with at least one valid functional alpha and beta chain, and kept only a single chain for each cell where there were multiple chains. We performed the analysis both on the alpha and beta chain separately, as well as on paired alpha and beta chain, in each instance requiring exact matching of the CDR3 amino acid sequences.

---

**Supplementary Figure 1:**
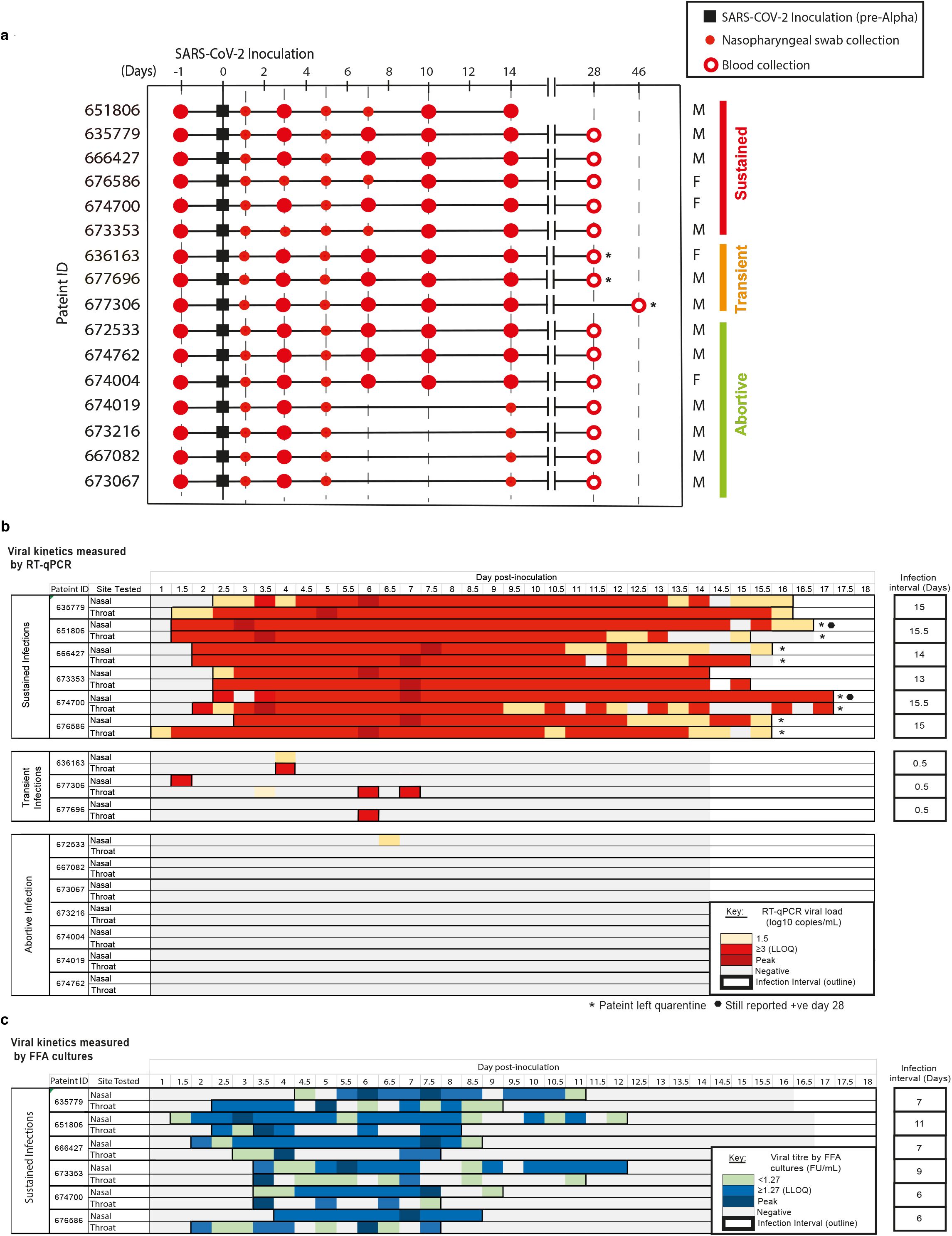
Overview of Single Cell Human SARS-CoV-2 Challenge Study cohort. **(a)** Timeline of the samples collected from each of the 16 participants enrolled in our study. Sample collections are shown relative to the date of SARS-CoV-2 inoculation (day 0). Samples are shown by infection group (sustained, transient and abortive), with their age in years (yrs) and sex (self-identified). *Indicates participants who were either vaccinated (636163) or reported to have developed an community infection, before or immediately after blood samples were taken on day 28 (677696 and 677306). See sample collection section in the methods for more details. Longitudinal measures of nasal and pharyngeal (throat) viral kinetics from swabs. Shown for each participant as measured via **(b)** RT-qPCR and **(c)** quantitative culture by focus forming assay (FFA). Patients were identified as testing positive if they had at least one RT-qPCR test where the viral load was able to be quantified (≥lower limit of quantification (LLOQ)). Six participants were seen to present multiple, sequential, positive RT-qPCR results and were classified as having a sustained infection. Three participants were seen to have standalone positive results and were classified having transient infections. Seven participants never presented a single RT-qPCR test result **≥** LLOQ and these were classified as abortive infections. FFA tests were only performed for patients identified as having sustained infections, so there is no data for participants with transient or abortive infections. Infection intervals for each participant were calculated based on the first and last across the nose and throat, where positive tests below the LLOQ were counted if they occurred < 2 days of a quantifiable (≥ LLOQ) test result. *Indicated where the patient was discharged from quarantine prior to testing negative. The black octagon highlights patients that were still reporting positive results at day 28 post-inoculation.

**Supplementary Figure 2:**
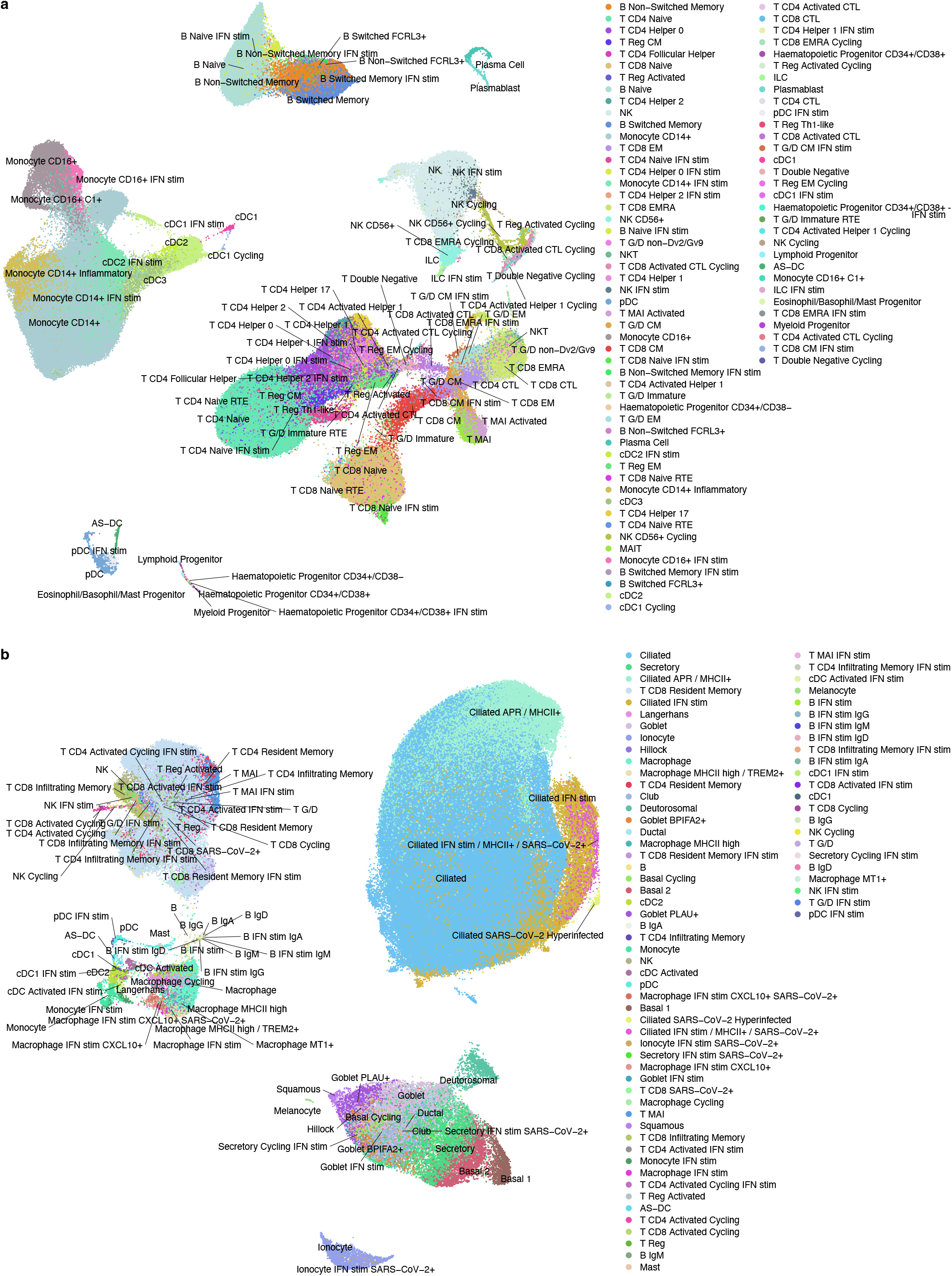
All identified and annotated cells. (**a**) UMAP of all PBMCs, color-coded and labeled by detailed cell state annotation. Subsets of B cells with differential immunoglobulin chain usage are not shown in full detail for clarity. (**b**) UMAP of all nasopharyngeal cells, color-coded and labeled by detailed cell state annotation.

**Supplementary Figure 3:**
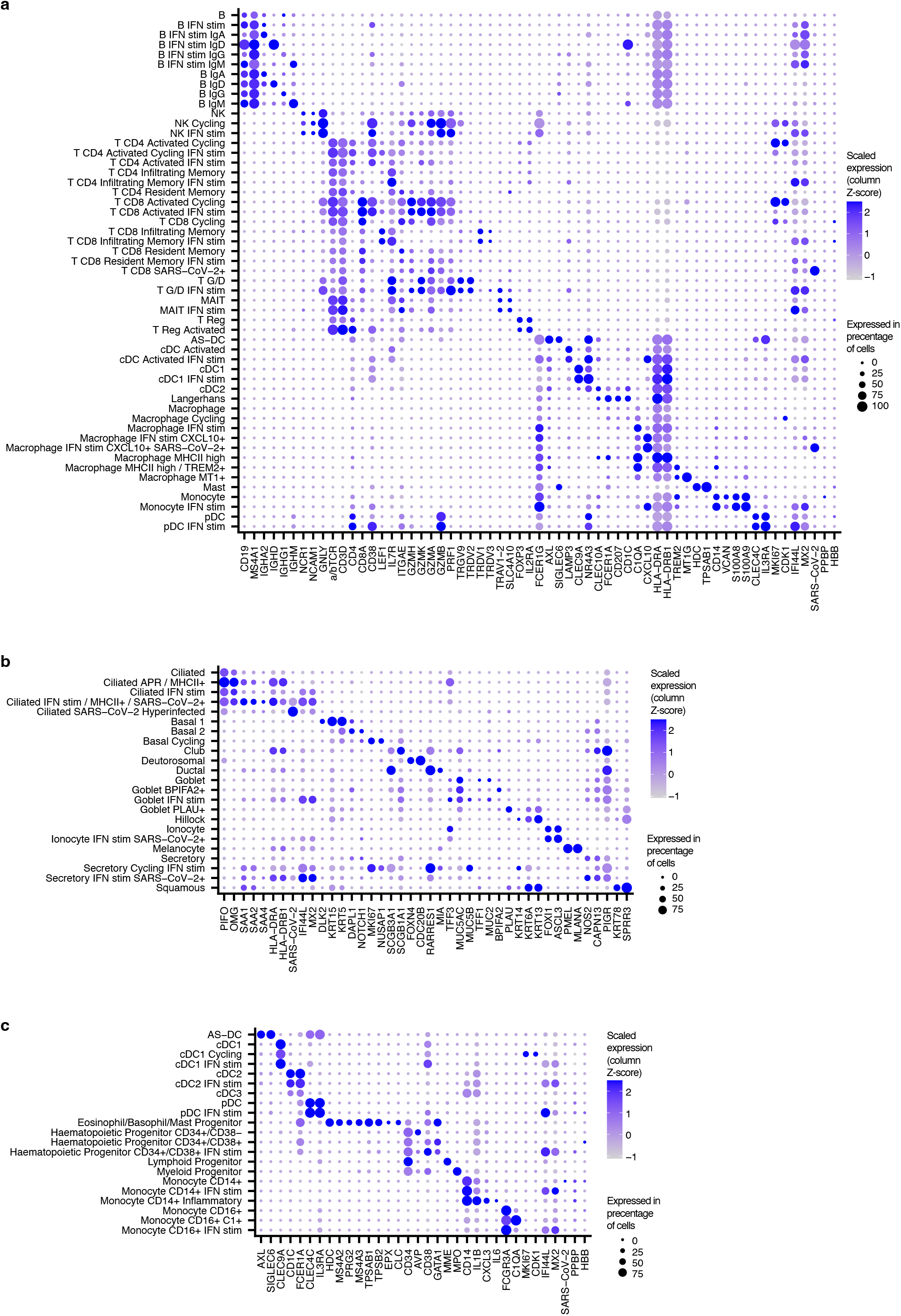
Marker gene expression used for annotation. Marker gene expression of cell states annotated in (**a**) nasopharyngeal immune cells, (**b**) nasopharyngeal epithelial cells, (**c**) myeloid and progenitor PBMCs.

**Supplementary Figure 4:**
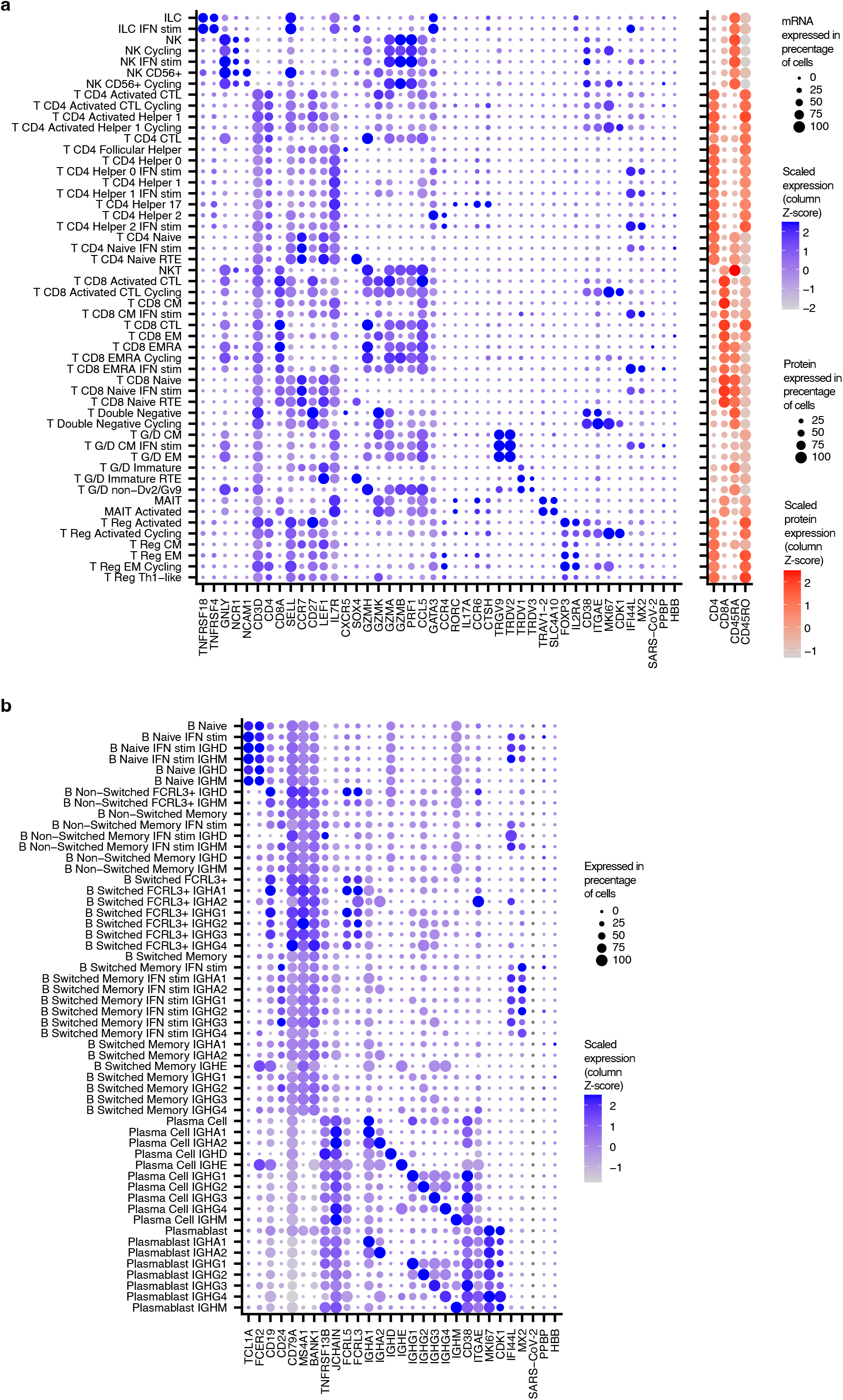
Marker gene expression used for annotation of PBMCs. Marker gene expression of cell states annotated in (**a**) T, NK and ILC cells in PBMCs, (**b**) B cells in PBMCs.

**Supplementary Figure 5:**
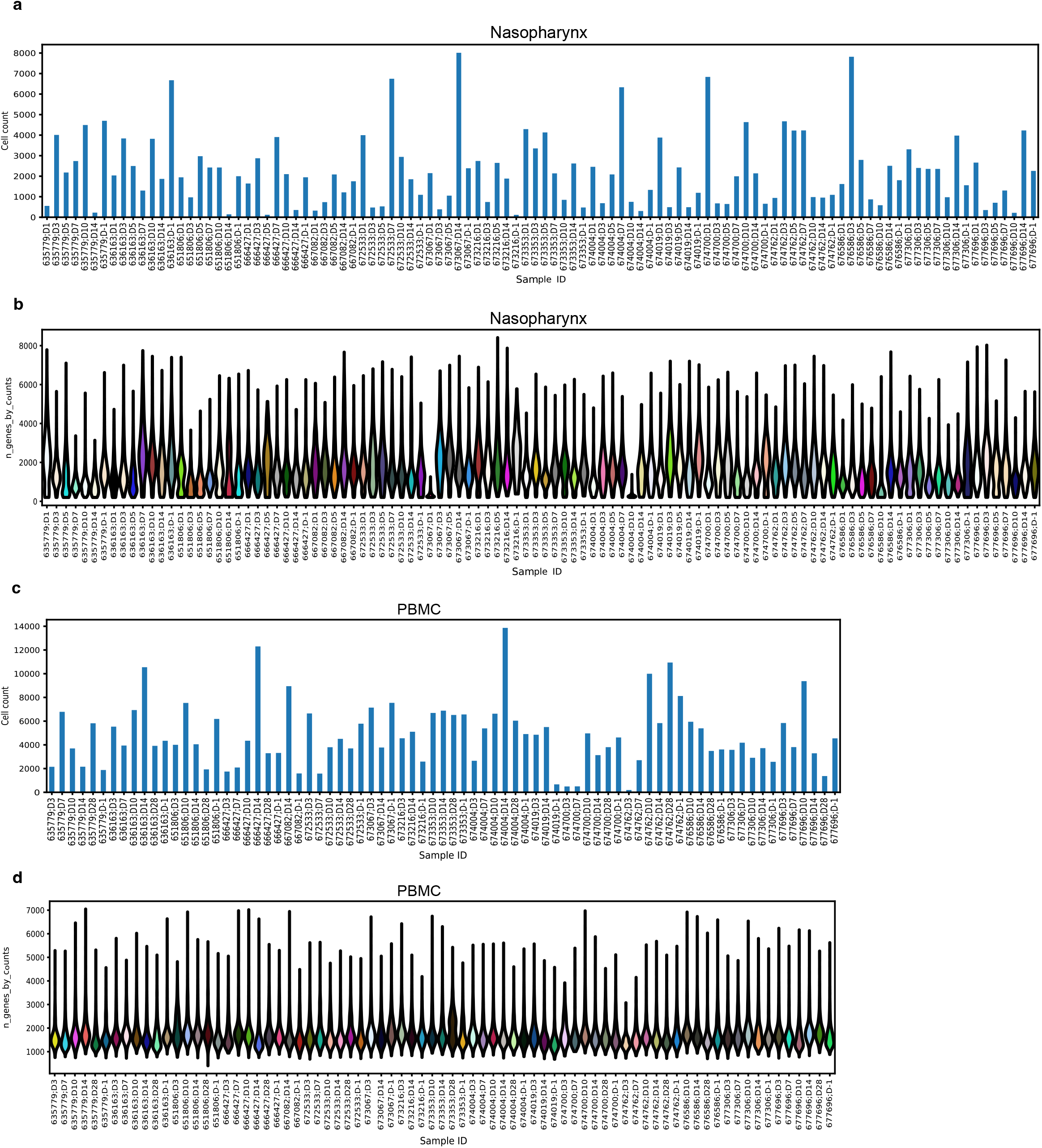
Quality control metrics of single cell RNA-seq data. Calculate quality control metrics for nasopharyngeal swabs showing **(a)** cells per sample and **(b)** gene reads per cell per sample. **(c)** and **(d)** show the same respectively for PBMCs.

**Supplementary Figure 6:**
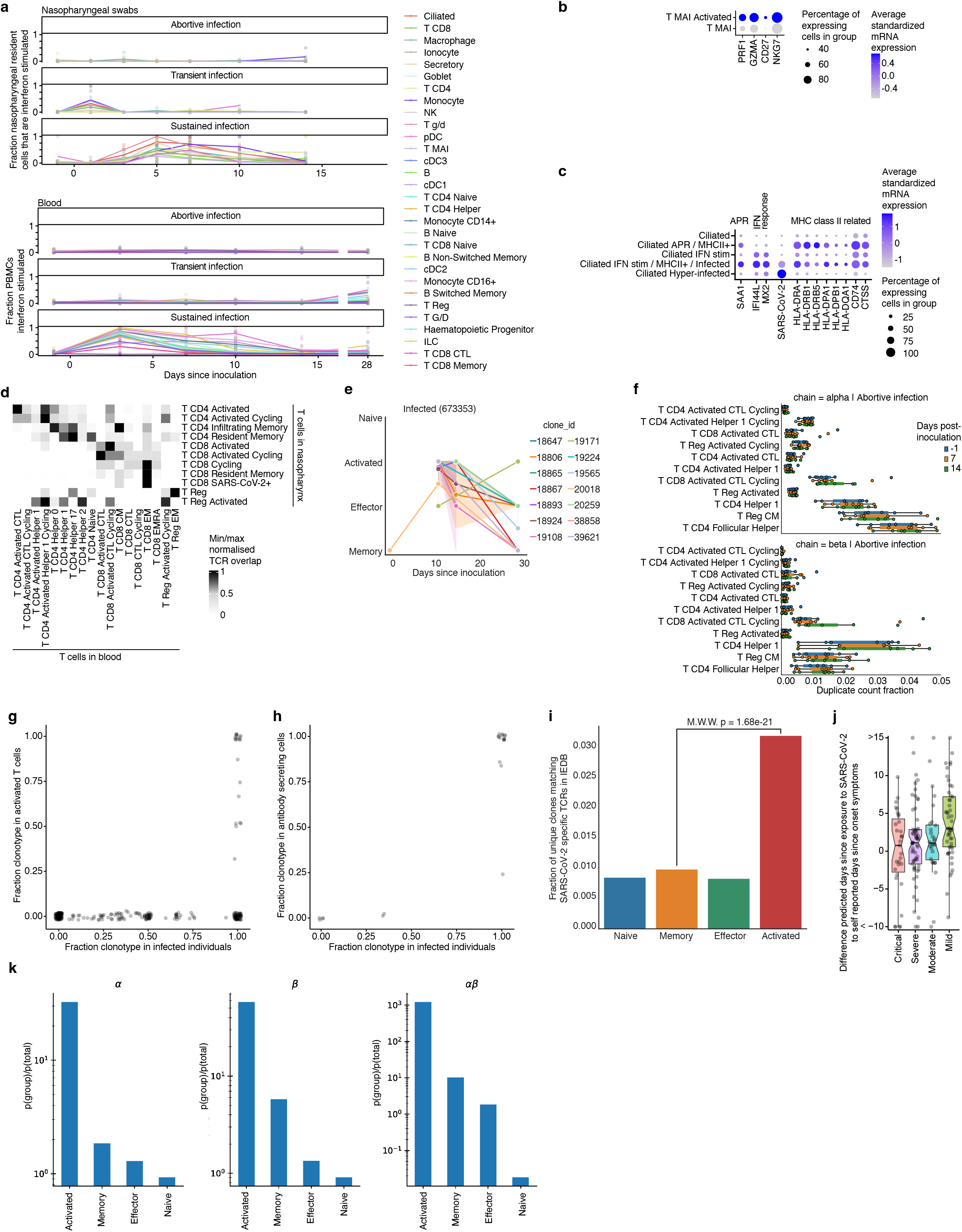
Temporal response states and activated T cells. (**a**) Line plot showing the mean proportions of interferon stimulated cells over time since inoculation within cell types with a distinct and annotated cluster of interferon stimulated cells. (**b**) Marker gene expression of activated MAIT cells. (**c**) Marker gene expression of response states observed in ciliated cells. (**d**) TCR repertoire overlap of nasopharyngeal and circulating conventional T cells. We only considered the beta TCR chain to identify overlapping T cells to include T cells without a TRA sequence detected. (**e**) Memory analysis in an individual with sustained SARS-CoV-2 infection. Unique TCR clones are distinguished by color and numbered with their clone_id identifier. A shaded area is drawn when the same clone appeared with several distinct cell type labels, and the size of the shaded area informs their relative ratios. (**f**) TCR bulk data with matched single cell labels as in Fig. 3g, but showing data on abortive infections for activated and other T cells. No particular changes are observed across the three time points sampled. (**g**) The fraction of activated T cells that participate in TCR clonotype groups versus the fraction of cells in each group that originate from participants with sustained infections. (**h**) Scatterplot as in (g), but showing BCR clonotype groups and the fraction of antibody secreting B cells instead of activated T cells. (**i**) Fraction of unique clones matching SARS-CoV-2 entries in IEDB across all T cell clones within that broad T cell compartment. Significance level after Whitney-Mann testing shown for Activated vs Memory T cells (putative SARS-CoV-2 fraction 3.75 times higher in Activated T cells, p = 1.68 ∗ 10−21) (**j**) Difference between predicted time since viral exposure and reported time since onset of symptoms, split by reported severity. In all box plots, the central line and the notch are the median and its approximate 95% confidence interval, the box shows the interquartile range and the whiskers are extreme values upon removing outliers (**k**) Coincidence analysis of TCR sequence diversity restriction in phenotypic subsets. Fraction of clonotype pairs within each phenotypic cluster that share identical CDR3 amino acid sequences (but distinct nucleotide sequences) normalized by the same statistics calculated across all clonotypes, for alpha, beta, and both chains together. The ratio of within cluster versus overall sequence coincidence probabilities is a measure of the breadth of epitopes targeted by the different clonotypes within a cluster ^52^.

**Supplementary Figure 7:**
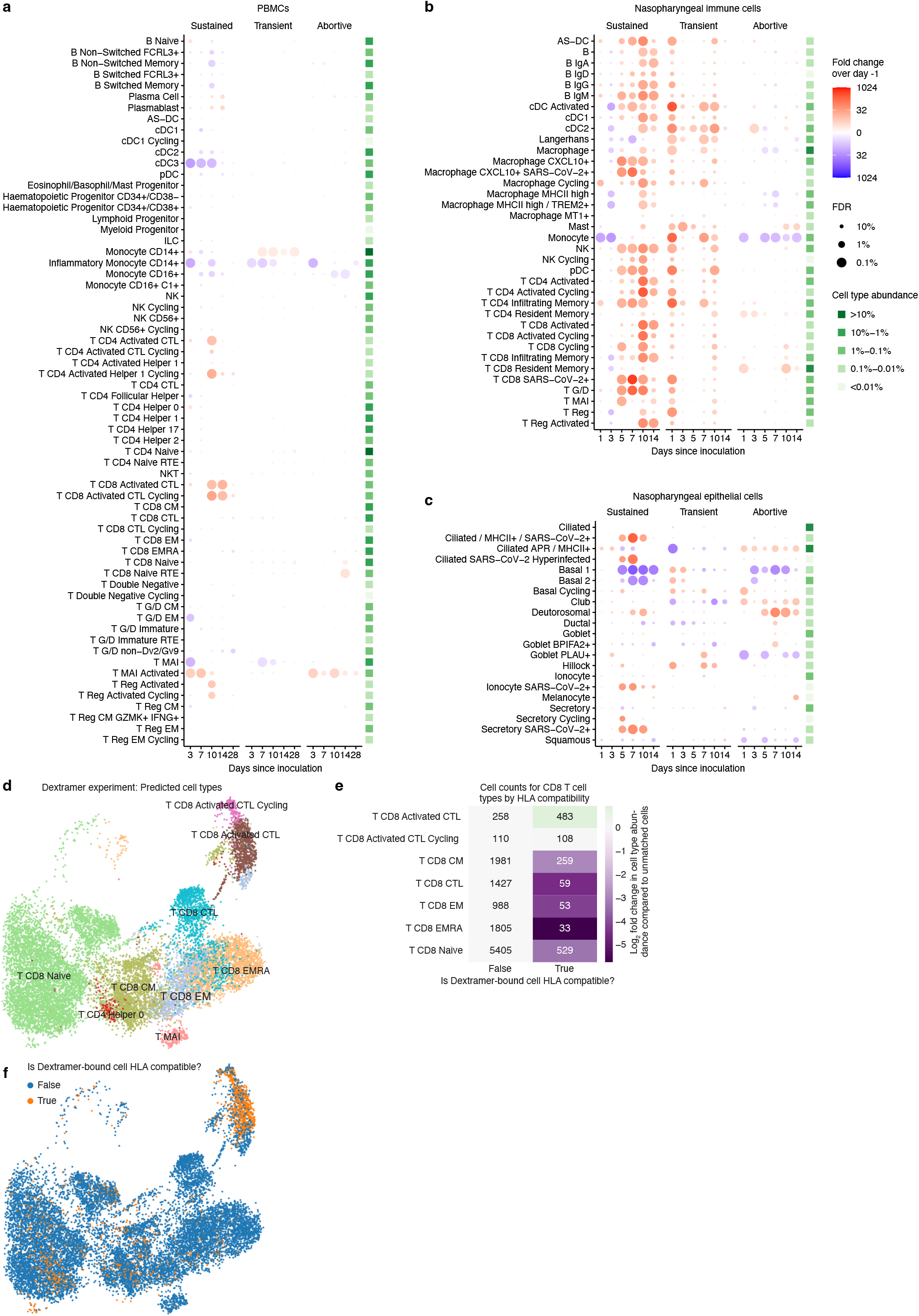
Detailed temporal dynamics in cell state abundances. (**a**) Fold changes in abundance of cell states in PBMCs. Detailed annotation of interferon stimulated subsets and immunoglobulin class specific cell states are not shown for clarity. Immune cell abundances were scaled to the total amount of detected PBMCs in every sample prior to calculating the fold changes over days since inoculation compared to pre-infection (day -1) by fitting a GLMM on scaled abundances. The mean cell type proportions over all cells and samples is shown in the green heatmap right of the dotplot to aid the interpretation of changes in cell type abundances. (**b**) Dotplot as in (a), but showing nasopharyngeal immune cells. Immune cell abundances were scaled to the total amount of detected epithelial cells in every sample. (**c**) Dotplot as in (a), but showing nasopharyngeal epithelial cells. **(d)** UMAP of all CD8+ T cells from the Dextramer assay, with cell types predicted by Celltypist model trained on previous PBMC data. Activated T cells form a distinct cluster. **(e)** Cell counts for CD8+ T cell types by HLA compatibility of donor with the highest-bound Dextramer. Only Activated T cells have positive log2 fold change for HLA-matched Dextramers. **(f)** UMAP as in (d), colored by HLA compatibility.

**Supplementary Figure 8:**
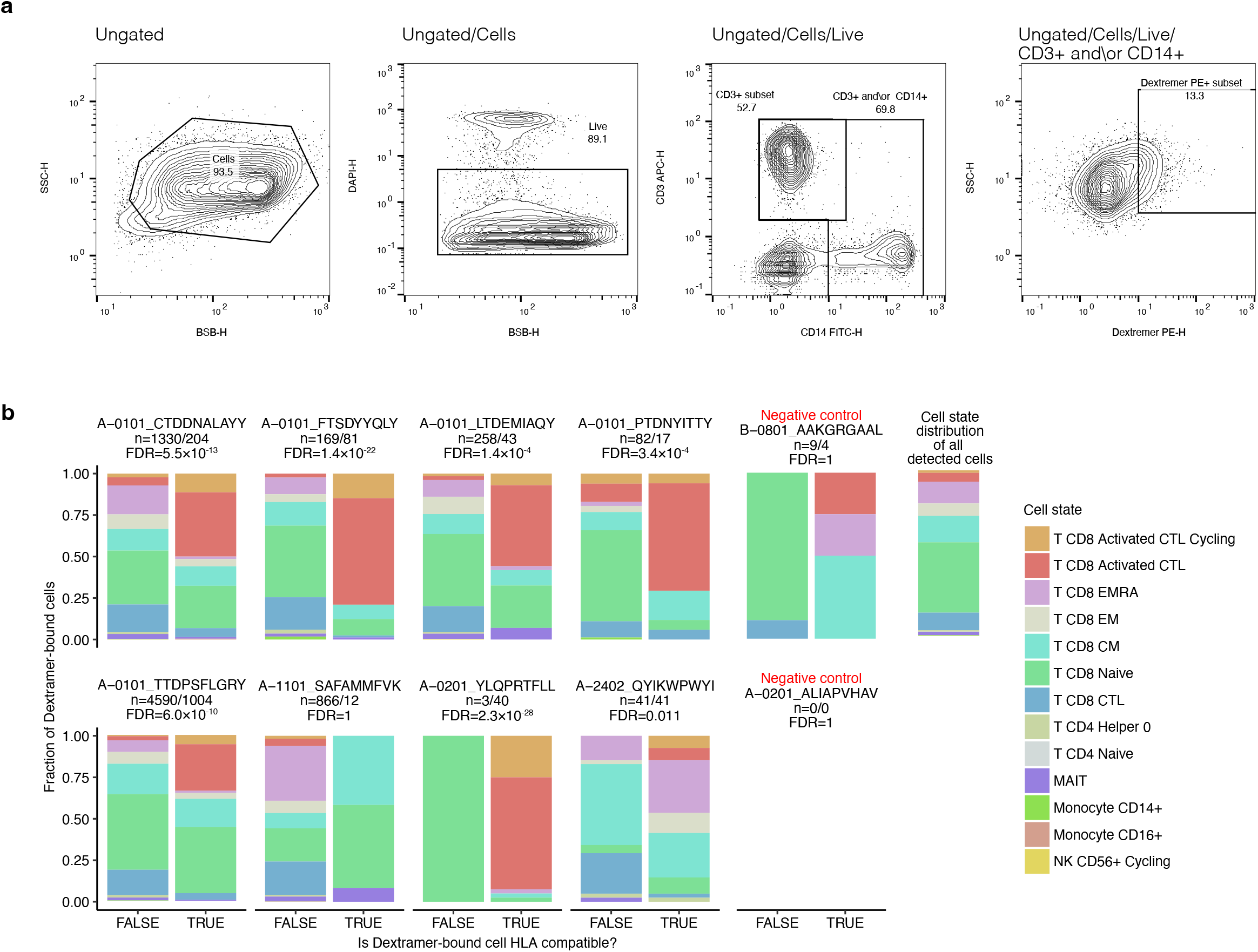
Validation of antigen-specific activated T cells. (**a**) Gating strategy used to enrich SARS-CoV-2 antigen specific T cells via MACSQuant Tyto cell sorting. Cells were sequentially stained with a multi-allele panel of dCODE dextramer-PE complexes, with the addition of anti-human CD3-APC and CD14-FITC FACS antibodies as references to help us identify T cell specific binding. Debris and cell aggregates were gated out first using BSB-H (backscatter blue laser-height) SSC-H (side scatter-height). From the cells, DAPI+ dead cells were excluded. T cells (CD3+) and monocytes (CD14+) were then gated for (CD3+ and\or CD14+ population) and the sort gate defined from this population as all PE-dCODE Dextramer® positive cells. This lenient sorting strategy was decided upon in order to collect enough cells for 10X 5’ single cell analysis downstream and to ensure we were capturing all SARS-CoV-2 antigen specific cells. Any non-specific binding (e.g. to monocytes) and background noise could then be removed computationally. (**b**) Calculated quality control metrics for PE-dCODE Dextramer® positive sequenced cells showing gene reads per cell per sample. (**c**) Proportions of activated T cells bound to Dextramers loaded with selected SARS-CoV-2 antigens. The total amount of bound cells to each Dextramer is shown, color-coded by predicted cell state. If barcodes from several Dextramers were detected to be bound to the same cell, we only selected the Dextramer with the highest signal as bound. As a control to separate background and real binding, cells are separated based on the HLA haplotype compatibility with the tested Dextramer. Only Dextramers with at least 10 HLA matched bound cells are shown. FDR corrected p values were determined by a Fisher-exact test comparing the proportion of HLA matched activated T cells in the Dextramer bound cells to the proportion of unbound HLA matched activated T cells. N represents the number of cells in each bar. The right-most bar represents the overall distribution of cell types across all Dextramer experiments.

## Notes

### Clinical Trial

NCT04865237

### Author Declarations

The UK Health Research Authority - Ad Hoc Specialist Ethics Committee gave ethical approval for this work (reference: 20/UK/2001 and 20/UK/0002).

